# Cumulative disease-course phenotypes in Crohn’s disease: a retrospective single-centre FAMD clustering study

**DOI:** 10.64898/2026.09.22.26363649

**Authors:** B. Caridi, C. Amoroso, L. Pozzi, D. Noviello, A. Arcidiacono, L. Baldari, L. Boni, M. Prati, S. Costa, B. Oreggia, F. Facciotti, F. Caprioli

## Abstract

**Background and aims:** Crohn’s disease (CD) is a highly heterogeneous disease. Patients with similar characteristics at baseline may end up on very different therapeutic and surgical paths. We aimed to identify disease-course trajectory phenotypes from a mix of clinical, treatment, surgical and comorbidity variables, and evaluate whether information already available at diagnosis could anticipate which trajectory a patient would follow.

**Methods:** We analysed a retrospective cohort of 98 unique CD patients enrolled at the IRCCS Ca’ Granda Policlinico Hospital, Milan. Factor analysis of mixed data (FAMD) was applied to 30 disease-course variables spanning demographic and Montreal descriptors, extra-intestinal manifestation burden, individual advanced-therapy drug exposures, surgical burden and comorbidity/symptom burden; clusters were then ranked by treatment burden using a line-weighted therapy score built separately from drug line positions and therapeutic-class weights. Clustering itself was carried out in the retained FAMD coordinate space, and cluster robustness was checked with silhouette metrics and bootstrap Jaccard stability. Baseline-associated variables were assessed separately, while advanced-therapy episodes were reconstructed from the therapy timeline.

**Results:** Three trajectory phenotypes emerged: C1 (n=56), C2 (n=31) and C3 (n=11). C3 was characterised by younger age at diagnosis, a heavier extra-intestinal manifestation burden, more intensive and prolonged therapy, greater biologic exposure and a higher cumulative number of surgeries, with comorbidity burden rising across the gradient. Montreal behaviour and comorbidity burden emerged as the strongest baseline correlate of trajectory membership, ahead of age at diagnosis and extra-intestinal manifestation burden; penetrating (B3) behaviour, notably, was actually more common in C1 than in C3. A model built on the diagnosis-available variables independently linked to trajectory membership (age at diagnosis, disease location and Montreal behaviour) showed moderate discrimination for the C3 trajectory (area under the curve, AUC=0.80, 95% CI 0.67-0.93). A sensitivity analysis that also added extra-intestinal manifestation count and comorbidity burden, whose timing relative to diagnosis we could not confirm, pushed discrimination to AUC=0.90.

**Conclusions:** FAMD-space clustering identified stable, clinically interpretable CD-course trajectory phenotypes, including a small, high-burden C3 subgroup. This C3 trajectory, small, early-onset and treatment-refractory, shares clinical features that overlap with the difficult-to-treat CD phenotype, though C3 was derived by unsupervised clustering rather than validated against that framework’s own criteria; variables available at or near diagnosis showed exploratory discrimination for the high-burden C3 trajectory, though external validation and careful timing verification are needed before any clinical use.

## Introduction

Crohn’s disease (CD) is a chronic, relapsing inflammatory disorder of the gastrointestinal tract characterised by substantial clinical heterogeneity [1]. Patients differ in age at diagnosis, inflammatory burden, disease location, behaviour, extra-intestinal manifestations, comorbidity burden, need for advanced therapy and surgical history [1]. Such heterogeneity extends to treatment response itself: loss of response to biologic therapy is common but highly variable, with meta-analytic estimates of anti-TNF loss of response ranging from 8% to 71% across studies (pooled incidence 33%, 95% CI 29-38, at one year) [2], and comparable variability reported for newer agents such as vedolizumab [3]. Real-world treatment approach for CD is itself heterogeneous, with population-based data showing substantial geographic variation in which pharmacological therapies are prescribed and in the outcomes achieved [4]. Because the advanced-therapy landscape available to clinicians has also expanded considerably over the past two decades, disease-course trajectories inevitably reflect both individual patient biology and the historical period in which each patient was treated, a dimension that baseline, single-time-point classification cannot capture.

The Montreal classification is the current reference standard for CD phenotyping [5]. It stratifies patients by age at diagnosis (A1 ≤16 years, A2 17-40 years, A3 >40 years), disease location (L1 ileal, L2 colonic, L3 ileocolonic, with L4 upper gastrointestinal disease recorded as a modifier) and behaviour (B1 non-stricturing/non-penetrating, B2 stricturing, B3 penetrating, with a perianal modifier) [5]. A paediatric adaptation, the Paris classification, extends this framework with growth-impairment and age-of-onset categories relevant to childhood-onset disease [6]. Both systems remain the standard for phenotypic description in clinical practice and trials, but they classify a patient at a single time point, typically at or near diagnosis, and do not, by design, capture how a patient’s disease course evolves therapeutically and surgically over years of follow-up.

That limitation has fuelled growing interest in data-driven, computational approaches to phenotyping in inflammatory bowel disease, spanning genomic, transcriptomic and clinical machine-learning applications [7]. Unsupervised clustering, in particular, has begun to reveal patient subgroups that single-time-point classification alone would miss: a recent population-based study, for example, used unsupervised machine learning to identify distinct multimorbidity clusters among patients with inflammatory bowel disease, with direct implications for tailored multidisciplinary care [8]. Few studies, however, have applied this approach specifically to longitudinal CD-course burden, integrating cumulative treatment, surgical and comorbidity history into a single data-driven phenotype, an approach explored here. Most work instead integrates genomic or single-domain clinical data, as in the transcriptomic clustering used to define molecular IBD subtypes [9]. A small subset of patients with inflammatory bowel disease experience persistent disease activity despite sequential exposure to multiple therapeutic classes, a phenomenon recently given a formal definition by international consensus, i.e., “difficult-to-treat” inflammatory bowel disease [10]. Identifying these patients, and ideally anticipating them before their disease course becomes entrenched, is a clinical priority underscored by real-world cohort data on this population’s prevalence, features and outcomes [11].

Existing trajectory-based work in CD has generally clustered patients on a single, repeatedly measured marker of disease activity or treatment response, rather than on the broader mix of treatment, surgical and comorbidity history used here. Longitudinal faecal calprotectin and C-reactive protein have been used this way in two large European cohorts [12,13], and a similar approach applied to C-reactive protein identified infliximab-response subtypes in paediatric CD [14]. A separate strand of research, mostly post hoc analyses of randomised trials, has modelled response trajectories to individual advanced therapies such as filgotinib in ulcerative colitis [15]; a recent review argues this kind of trajectory-based precision medicine is where the field is heading [16]. Symptom clusters and multi-omics stratification tackle the same heterogeneity from different angles: the former predicts healthcare utilisation from patient-reported symptoms [17], the latter groups patients by molecular subtype [18]. What sets the present study apart is scope rather than method: rather than tracking one marker or one drug’s response over time, we cluster on the cumulative clinical, therapeutic, surgical and comorbidity history of the whole disease course, captured at a later retrospective time point.

We designed the present study around two related but distinct tasks. First, we aimed to identify data-driven disease-course trajectory phenotypes using variables that summarise longitudinal clinical burden, treatment exposure, surgery and comorbidity. Second, once those trajectories were defined, we asked whether information available at or near diagnosis could classify patients into the resulting phenotypes, or flag those at risk of the most severe trajectory.

## Methods

### Human Subjects

This single-centre study was conducted at Fondazione IRCCS Ca’ Granda Ospedale Maggiore Policlinico (Milan, Italy). Ethical approval for this study was obtained from the CE Milano Area 2, number 566_2015 quinquies, 557_2018, 128_2018bis, 4553_2024. Patient disease-course history was obtained from patient clinical records, and cumulative clinical, therapeutic and surgical history was summarised on a single de-identified Excel workbook, organised into labelled sheets (FAMD variables, clinical and surgical data, therapy timeline and scores, and a variable legend). Patient names are not included in this workbook; a separate, internal-only file that is not part of any shared or published dataset links the study ID to identifying information for internal verification purposes only. Input data quality is documented through automated checks for duplicate patient records and an internally logged data-quality-flags sheet. Patients were ascertained through the surgical/IBD referral pathway at Policlinico of Milan; consistent with this ascertainment, all patients had at least one recorded Crohn-related surgery or biopsy, and the cohort is therefore enriched for surgically-referred disease relative to general CD populations (**Table 1**). Of the 130 patients initially identified through this pathway, 32 were excluded for insufficient documentation or the lack of a recorded Crohn-related surgery or biopsy, leaving the final cohort of 98 patients (94 with recorded surgery, 4 by biopsy alone). The surgical time-to-event endpoint is defined as time to surgery recorded at Policlinico of Milan, not first lifetime surgery. The characteristics of enrolled patients (n = 98) are reported in **Table 1**. Continuous variables are presented as median and interquartile range (IQR); categorical data are presented as counts and percentages.

**Table 1.** Baseline demographic, disease-phenotype and treatment-burden characteristics by cluster.

| Characteristic | Overall<br>(N=98) | C1 (n=56) | C2 (n=31) | C3 (n=11) | P value | P adj (test; effect size) |
| --- | --- | --- | --- | --- | --- | --- |
| Demographics |  |  |  |  |  |  |
| Age at diagnosis, years,<br>median (IQR) | 29.3 (20.6-42.2) | 27.7 (20.3-39.1) | 39.5 (28.6-46.5) | 19.9 (18.5-24.5) | <0.001 | 0.001 (KW;<br>epsilon2=0.157) |
| Current age, years, median<br>(IQR) | 51.5 (40.1-59.3) | 48.0 (37.7-57.4) | 59.1 (51.3-69.5) | 50.0 (37.1-54.9) | 0.001 | 0.003 (KW;<br>epsilon2=0.136) |
| Disease duration, years,<br>median (IQR) | 17.6 (11.3-24.3) | 14.3 (10.3-22.5) | 18.3 (14.3-24.3) | 29.3 (21.3-30.8) | 0.012 | 0.023 (KW;<br>epsilon2=0.091) |
| Female sex, n (%) | 50 (51.0%) | 25 (44.6%) | 20 (64.5%) | 5 (45.5%) | 0.196 | 0.219 (chi-sq;<br>CramersV=0.184) |
| Disease phenotype |  |  |  |  |  |  |
| Montreal age at diagnosis |  |  |  |  |  |  |
| A1 (<17 years) | 7 (7.1%) | 6 (10.7%) | 0 (0.0%) | 1 (9.1%) | 0.014 | 0.023 (chi-sq;<br>CramersV=0.254) |
| A2 (17-40 years) | 62 (63.3%) | 36 (64.3%) | 16 (51.6%) | 10 (90.9%) |  |  |
| A3 (>40 years) | 29 (29.6%) | 14 (25.0%) | 15 (48.4%) | 0 (0.0%) |  |  |
| Disease location |  |  |  |  |  |  |
| L1 ileal | 30 (30.6%) | 16 (28.6%) | 12 (38.7%) | 2 (18.2%) | 0.003 | 0.006 (chi-sq;<br>CramersV=0.358) |
| L2 colonic | 2 (2.0%) | 0 (0.0%) | 0 (0.0%) | 2 (18.2%) |  |  |
| L3 ileocolonic | 28 (28.6%) | 21 (37.5%) | 6 (19.4%) | 1 (9.1%) |  |  |
| Multiple locations | 36 (36.7%) | 18 (32.1%) | 13 (41.9%) | 5 (45.5%) |  |  |
| Unknown | 2 (2.0%) | 1 (1.8%) | 0 (0.0%) | 1 (9.1%) |  |  |
| Disease behaviour |  |  |  |  |  |  |
| B1 non-stricturing/non-penetrating | 7 (7.1%) | 5 (8.9%) | 1 (3.2%) | 1 (9.1%) | <0.001 | <0.001 (chi-sq;<br>CramersV=0.478) |
| B2 stricturing | 47 (48.0%) | 18 (32.1%) | 26 (83.9%) | 3 (27.3%) |  |  |
| B2B3 | 7 (7.1%) | 3 (5.4%) | 1 (3.2%) | 3 (27.3%) |  |  |
| B3 penetrating | 32 (32.7%) | 28 (50.0%) | 3 (9.7%) | 1 (9.1%) |  |  |
| Unknown | 5 (5.1%) | 2 (3.6%) | 0 (0.0%) | 3 (27.3%) |  |  |
| Smoking status |  |  |  |  |  |  |
| Ex-Smoker | 22 (22.4%) | 8 (14.3%) | 11 (35.5%) | 3 (27.3%) | 0.280 | 0.280 (chi-sq;<br>CramersV=0.269) |
| Heavy-Smoker | 2 (2.0%) | 2 (3.6%) | 0 (0.0%) | 0 (0.0%) |  |  |
| Light-Smoker | 8 (8.2%) | 5 (8.9%) | 2 (6.5%) | 1 (9.1%) |  |  |
| Mod-Smoker | 10 (10.2%) | 6 (10.7%) | 2 (6.5%) | 2 (18.2%) |  |  |
| Non-Smoker | 42 (42.9%) | 26 (46.4%) | 13 (41.9%) | 3 (27.3%) |  |  |
| Occ-Smoker | 5 (5.1%) | 3 (5.4%) | 0 (0.0%) | 2 (18.2%) |  |  |
| Unknown | 9 (9.2%) | 6 (10.7%) | 3 (9.7%) | 0 (0.0%) |  |  |
| Any extraintestinal manifestation, n (%) | 73 (74.5%) | 38 (67.9%) | 24 (77.4%) | 11 (100.0%) | 0.074 | 0.100 (chi-sq; CramersV=0.230) |
| Number of extraintestinal manifestations, median (IQR) | 3 (0-5) | 2 (0-3) | 4 (2-5) | 5 (3-8) | <0.001 | <0.001 (KW; epsilon2=0.170) |
| Associated pathologies |  |  |  |  |  |  |
| NO | 36 (36.7%) | 33 (58.9%) | 1 (3.2%) | 2 (18.2%) | <0.001 | <0.001 (chi-sq; CramersV=0.414) |
| Unknown | 3 (3.1%) | 3 (5.4%) | 0 (0.0%) | 0 (0.0%) |  |  |
| YES | 59 (60.2%) | 20 (35.7%) | 30 (96.8%) | 9 (81.8%) |  |  |
| Disease course and treatment burden |  |  |  |  |  |  |
| Any advanced therapy exposure, n (%) | 79 (80.6%) | 42 (75.0%) | 26 (83.9%) | 11 (100.0%) | 0.139 | 0.176 (chi-sq; CramersV=0.202) |
| Number of advanced therapies, median (IQR) | 1 (1-2) | 1 (1-2) | 1 (1-2) | 4 (3-6) | <0.001 | <0.001 (KW; epsilon2=0.243) |
| Number of non-biologic therapies, median (IQR) | 1 (1-2) | 1 (1-1) | 2 (1-2) | 2 (2-2) | <0.001 | <0.001 (KW; epsilon2=0.292) |
| Corticosteroid exposure, n (%) | 70 (71.4%) | 34 (60.7%) | 26 (83.9%) | 10 (90.9%) | 0.025 | 0.036 (chi-sq; CramersV=0.278) |
| Therapy score, median (IQR) | 8.5 (2.5-17.0) | 4.2 (2.5-11.1) | 11.0 (6.8-17.5) | 46.5 (31.5-66.8) | <0.001 | <0.001 (KW; epsilon2=0.364) |
| Any surgery, n (%) | 94 (95.9%) | 52 (92.9%) | 31 (100.0%) | 11 (100.0%) | 0.251 | 0.265 (chi-sq; CramersV=0.179) |
| Number of surgeries, median (IQR) | 1 (1-3) | 1 (1-2) | 1 (1-2) | 3 (1-4) | 0.024 | 0.036 (KW; epsilon2=0.077) |
| Years from diagnosis to first biologic, median (IQR) | 5.3 (2.2-11.6) | 4.0 (1.6-9.0) | 7.5 (3.1-15.8) | 10.0 (3.7-15.3) | 0.158 | 0.188 (KW; epsilon2=0.050) |

*P value is the raw test P value; P adj is the Benjamini-Hochberg-adjusted P value across this table's family of parallel univariate comparisons (Kruskal-Wallis for continuous variables, chi-square with simulated P values for categorical variables); effect size is epsilon-squared or Cramér's V as noted. For multi-level categorical variables the P values and effect size are reported once, on the variable's first row. KW, Kruskal-Wallis; chi-sq, chi-square; epsilon2, epsilon-squared; CramersV, Cramér's V.*

### Trajectory Clustering

Throughout this manuscript, ‘trajectory’ refers to the disease-course phenotype reconstructed from each patient’s cumulative clinical, therapeutic and surgical history up to the data-cutoff, not to a repeated-measures or time-series model of disease activity over time; no variable in the active set is a longitudinal measurement taken at multiple time points. The final disease-course variable set included demographic/Montreal descriptors, extra-intestinal manifestation (EIM) burden, age at diagnosis, disease duration, individual advanced-therapy drug exposures, surgery burden and comorbidity/symptom burden. Categorical active variables were Gender, Behaviour, Disease_Site, Smoke_Rate, Montreal_A and Associated_Pathologies. Sex (Gender) was included as a categorical active variable for completeness, but contributed minimally to the three retained clustering dimensions (0.25%, 0.63% and 0.14% of Dimension 1, 2 and 3 variance respectively; about 1.0% combined). Numerical active variables were EIMs, Age_at_Diagnosis, Disease_Duration_Years, IFX, ADA, VDZ, UTK, UPA, Risa, Corticosteroids, MES, AZA, MTX, NO_Surgeries, NO_Pathologies, Weight_Loss, Diarrhea, Abdominal_Pain, Bowel_Obstruction, Fistulizing, Autoimmune_Diseases, Osteoporosis, Hypothyroidism and Infectious_Diseases. Drug codes were IFX (infliximab), ADA (adalimumab), VDZ (vedolizumab), UTK (ustekinumab), UPA (upadacitinib), Risa (risankizumab), MES (mesalamine), AZA (azathioprine) and MTX (methotrexate). Associated_Pathologies was a three-level categorical indicator (NO, YES, Unknown) of whether a patient had at least one documented comorbidity from this fixed tracked panel (autoimmune disease, infectious disease, metabolic disease, osteoporosis, hypothyroidism, anxiety/depressive disorder), four of which (autoimmune disease, infectious disease, osteoporosis and hypothyroidism) were also analysed as separate active variables; EIMs was a cumulative count of documented extra-intestinal manifestations, a conceptually distinct, IBD-specific category not overlapping with these comorbidities. Therapy_Score and its component summary variables (num_biologics, num_non_biologic_therapies, biologic_exposure_any, steroid_exposure) were excluded from FAMD as redundant aggregates of the individual drug-exposure variables already included, though Therapy_Score was retained to rank the resulting clusters by treatment burden. Montreal_A was derived from Age_at_Diagnosis using standard cut points: A1 ≤16 years, A2 17-40 years and A3 >40 years [5]. Current_Age was excluded to avoid redundancy with age at diagnosis and disease duration. Behaviour was kept at its five raw source-data levels (B1, B2, B2B3, B3, Unknown), where B2B3 denotes coexisting stricturing and penetrating behaviour. Before FAMD, eight binary comorbidity/symptom indicators (weight loss, diarrhoea, abdominal pain, bowel obstruction, autoimmune disease, osteoporosis, hypothyroidism, infectious disease) each had 3-4 missing values out of 98 patients, imputed with the variable mean. No categorical active variable needed mode imputation. The ‘Unknown’ categories kept in **Table 1** (Behaviour, disease location, smoking status, associated pathologies) are a distinct, explicitly recorded clinical category, not missing data that happened to be imputed away.

Given the combination of continuous and categorical measures in the disease-course variable set, dimensionality reduction was performed using Factor Analysis of Mixed Data (FAMD) [19], an extension of principal component analysis to mixed-type data. FAMD was computed with an initial number of retained dimensions (ncp) of 10; three dimensions were carried forward into clustering, chosen from exploratory comparisons of alternative dimension counts and cluster numbers (not shown) on the basis of bootstrap Jaccard stability and silhouette width. K-means clustering (stats::kmeans, nstart=50, iter.max=200, fixed seed of 42 for reproducibility) was applied to patient coordinates on the retained FAMD dimensions. Cluster labels were assigned by increasing mean Therapy_Score (C1 lowest, C3 highest). All analyses used R 4.5.2, with FactoMineR (2.15) for FAMD, glmnet (5.0) and pROC (1.19.0.1) for the ridge logistic models, geepack (1.3.13) for the clustered GEE comparisons, and logistf (1.26.1) for Firth logistic regression.

### Cluster Validation

Cluster validation included average and per-cluster silhouette [20], bootstrap Jaccard stability and comparison with alternative clustering solutions. The final clustering used k-means on the retained FAMD coordinates with multiple starts. Jaccard values above 0.50 were interpreted as supportive of stable clusters in this exploratory setting.

### Line-Weighted Therapy Score

Therapy burden was summarised using a line-weighted Therapy_Score, constructed for each patient across the set of tracked drugs. Each drug contributed zero to the score if never used, or its line position (or last observed line position) within the patient’s treatment sequence otherwise. The operational formula was: Therapy_Score = sum_i(Drug_i x Weight_i).

Each drug variable was coded 0 if never used, or as its line position (or last observed line position) within the patient’s treatment sequence if used, then multiplied by a class weight: infliximab, adalimumab, vedolizumab and ustekinumab (the legacy advanced biologics in this cohort) were weighted 1.0; upadacitinib and risankizumab (the newer advanced small-molecule/biologic agents) were weighted 1.5; and corticosteroids, mesalamine, azathioprine and methotrexate (conventional or steroid therapy) were weighted 0.5. Therapy_Score is the sum of these weighted line-position values across all tracked drugs for a given patient.

### Advanced-Therapy Analyses

Advanced-therapy episodes were reconstructed from the therapy timeline recorded in the input workbook, which was compiled patient-by-patient from each patient’s clinical chart directly into structured fields. For each episode, drug, line position, start/end year, duration where available, years since EU CD approval, treatment era and stop reason were derived. Documented failure was defined as primary failure, loss of response or side effect.

Drug names and abbreviations were standardised before analysis. Treatment era was defined relative to EU CD approval as pre-approval, early (0-4 years post-approval), established (5-9 years) or mature (≥10 years). Episode-level analyses included drug-line heatmaps, line-of-therapy position, duration distributions, stop-reason distributions, era-stratified failure rates and the number of advanced agents available at each patient’s year of diagnosis.

These episodes are not independent within a patient, so cluster comparisons at the episode level (treatment era, documented failure) were additionally modelled with a generalized estimating equation (GEE; geepack, binomial family, logit link, exchangeable working correlation, clustered by patient ID) alongside the naive chi-square test that treats episodes as independent; the naive chi-square P values are given in **Supplementary Tables 6 and 8**, and the main text reports the GEE estimate as the more conservative, non-independence-adjusted reference.

### Time-to-Event and Surgery Analyses

Kaplan-Meier analyses were used for time to first recorded biologic/advanced therapy and time to Policlinico of Milan surgery. Patients without the event were censored at recorded disease duration. Curves were compared by log-rank tests using survdiff. Surgical burden was also summarised as i) number of surgeries, ii) any surgery and iii) multiple surgery at patient level.

### Baseline Prediction

Random-forest models and a leave-one-out cross-validated (LOOCV) ridge logistic regression combining the strongest candidate predictors were used as internal exploratory audits rather than externally validated prediction tools. Random forests used inverse class-frequency weighting to reduce majority-class dominance, with cross-validated results interpreted only as an internal signal check.

Diagnosis-available candidate predictors were analysed after cluster construction. Candidate predictors were Age_at_Diagnosis, EIMs, Gender, Disease_Site, Behaviour, Smoke_Rate, Associated_Pathologies and Montreal_A. Therapy_Score, biologic counts, surgery and disease duration were excluded from baseline prediction because they encode future disease courses. Univariate associations were assessed using Kruskal-Wallis tests for continuous variables and chi-square tests with simulated p-values for categorical variables. Multiclass baseline classification used internally cross-validated multinomial models. Class-weighted models used inverse class-frequency weights within each training fold. Firth logistic C3-vs-rest screening was used to explore the small C3 class and reduce separation bias [21,22]. Age_at_Diagnosis, Disease_Site and Behaviour, the three variables kept in the primary model below, were also active FAMD clustering variables (see Trajectory Clustering above). Part of the model’s discriminative performance therefore simply reflects how well these variables already separate the clusters by construction, so we report it as an internal, exploratory re-description of the clustering rather than an independently validated prediction. For both models, discrimination is reported as the area under the curve (AUC) with a DeLong 95% confidence interval, and calibration as the intercept and slope from a logistic recalibration of the leave-one-out predicted probabilities against the observed outcome (perfect calibration: intercept 0, slope 1).

Candidate models additionally assessed whether Montreal_A, Disease_Site, Behaviour and Associated_Pathologies improved classification beyond age and EIM burden, individually and in combination. Montreal_A was interpreted cautiously because it is derived from age at diagnosis. Model performance was reported as cross-validated accuracy, balanced accuracy, macro-F1, class-specific sensitivity, precision and confusion matrices. C3 detection was summarised using threshold-based Firth logistic screening to show the trade-off between sensitivity and false positives, including screens combining the strongest individual predictors.

### Statistical Framing

Post-clustering descriptive analyses summarised continuous variables as median and interquartile range and categorical variables as count and percentage. Continuous variables were compared across three clusters using Kruskal-Wallis tests with epsilon-squared effect sizes; two-group sensitivity analyses used Wilcoxon rank-sum tests and rank-biserial effect sizes. Categorical variables used chi-square tests with simulated p-values when expected cell counts were small, and Cramér’s V was reported as an effect size. Episode-level comparisons (treatment era, documented failure), where a single patient can contribute multiple episodes, were additionally modelled with a generalized estimating equation clustered by patient (see Advanced-Therapy Analyses) to avoid overstating significance from within-patient correlation.

Six comorbidity variables (autoimmune disease, infectious disease, metabolic disease, osteoporosis, hypothyroidism and anxiety/depression) and seven presenting-symptom variables (fistulizing disease, weight loss, diarrhoea, abdominal pain, bowel obstruction, rectal bleeding and anaemia) were each compared across clusters using chi-square tests with simulated P values and summarised by cluster; the two variable groups formed separate Benjamini-Hochberg-adjusted test families. Smoking status was compared across clusters as a single chi-square test on the full categorical variable and was not part of either BH-adjusted family.

All prediction analyses were considered exploratory and internally validated only. As treatment and surgery contribute to the trajectory definition, associations between therapy burden and clusters were interpreted as phenotype-defining features, not independent predictors. Within each reported family of parallel univariate comparisons (**Table 1**; **Supplementary Table 2**; **Table 2**; the time-to-event/surgery family; and the comorbidity and presenting-symptom families), a Benjamini-Hochberg false-discovery-rate adjustment was applied and is reported alongside the raw P value as P adj; across these six families, 49 parallel comparisons were tested in total (19 in **Table 1**, 6 in **Supplementary Table 2**, 8 in **Table 2**, 3 in the time-to-event/surgery family, 6 for comorbidities and 7 for presenting symptoms); multivariable model coefficients and sensitivity-analysis model variants were not additionally adjusted, as they are not parallel independent hypothesis tests. A Benjamini-Hochberg-adjusted P value above 0.05 was not treated as evidence of no effect: in line with the argument that routine multiplicity correction can obscure clinically important associations in exploratory research [23], raw and adjusted P values are reported together throughout, and findings that lose significance after adjustment are interpreted in light of effect size, biological plausibility and consistency with external literature rather than discarded.

**Table 2.** Univariate candidate-predictor screening.

| Variable | Test | P adj (BH) | Effect size | C3-vs-rest OR (95% CI) |
| --- | --- | --- | --- | --- |
| Montreal behaviour | Chi-square with simulated P value | <0.001 | Cramér's V=0.478 | - |
| Associated comorbidities | Chi-square with simulated P value | <0.001 | Cramér's V=0.414 | - |
| Extra-intestinal manifestations | Kruskal-Wallis across clusters | <0.001 | epsilon2=0.170 | 1.55 (1.21–2.08) |
| Age at diagnosis | Kruskal-Wallis across clusters | 0.001 | epsilon2=0.157 | 0.93 (0.85–0.98) |
| Montreal location | Chi-square with simulated P value | 0.004 | Cramér's V=0.358 | - |
| Montreal age class | Chi-square with simulated P value | 0.016 | Cramér's V=0.254 | - |
| Sex | Chi-square with simulated P value | 0.224 | Cramér's V=0.184 | - |
| Smoking status | Chi-square with simulated P value | 0.276 | Cramér's V=0.269 | - |

## Results

### Three Disease-Course Trajectories were uncovered from Cumulative Clinical History

The final cohort comprised 98 patients (**Table 1**). To evaluate disease-course trajectory phenotypes independent of any single baseline classification, we performed a Factor Analysis of Mixed Data (FactoMineR::FAMD), taking into account 30 variables spanning demographic and Montreal descriptors, extra-intestinal manifestation burden, individual advanced-therapy drug exposures, surgical burden and comorbidity/symptom burden, collected from each patient’s cumulative clinical record **(Supplementary Figure 1**). FAMD explained 14.1% of variance on Dimension 1 and 8.0% on Dimension 2; the first three dimensions were retained for clustering, together capturing 28.2% of total variance **(Supplementary Figure 1A**). Dimension 1 was driven predominantly by treatment-exposure variables (corticosteroids, ustekinumab, infliximab and adalimumab), while Dimension 2 was driven predominantly by age- and comorbidity-related variables (number of associated pathologies, age at diagnosis, Montreal age class and presence of associated pathologies), as shown by the FAMD variable map and variable-contribution plots (**Supplementary Figure 1B, C)**. K-means clustering (nstart=50, iter.max=200, fixed seed) applied to patient coordinates on these three retained dimensions converged in 2 iterations (total within-cluster sum of squares = 616.83), yielding three clusters of 56 (C1), 31 (C2) and 11 (C3) patients (**Figure 1A**). The three identified clusters showed moderate-to-high bootstrap Jaccard stability (C1 0.76, C2 0.60, C3 0.67; **Figure 1B; Supplementary Table 1**) and an average silhouette width of 0.29 (**Supplementary Figure 1E**). Exploratory comparisons of alternative component counts and cluster numbers (not shown) supported k=3 as the most clinically interpretable solution, even though it was not the single top-ranked configuration by composite stability score: fewer clusters collapsed the C1/C2/C3 severity gradient central to this study’s phenotyping, while configurations with more clusters introduced poorly balanced groups. The within-cluster sum-of-squares elbow (**Supplementary Figure 1D**) declines smoothly without a single unambiguous inflection point, and per-cluster silhouette width (**Supplementary Figure 1E**) shows C1 achieving markedly better internal cohesion than C2 or C3, consistent with C1 being both the largest and most homogeneous trajectory.

**Figure 1.**
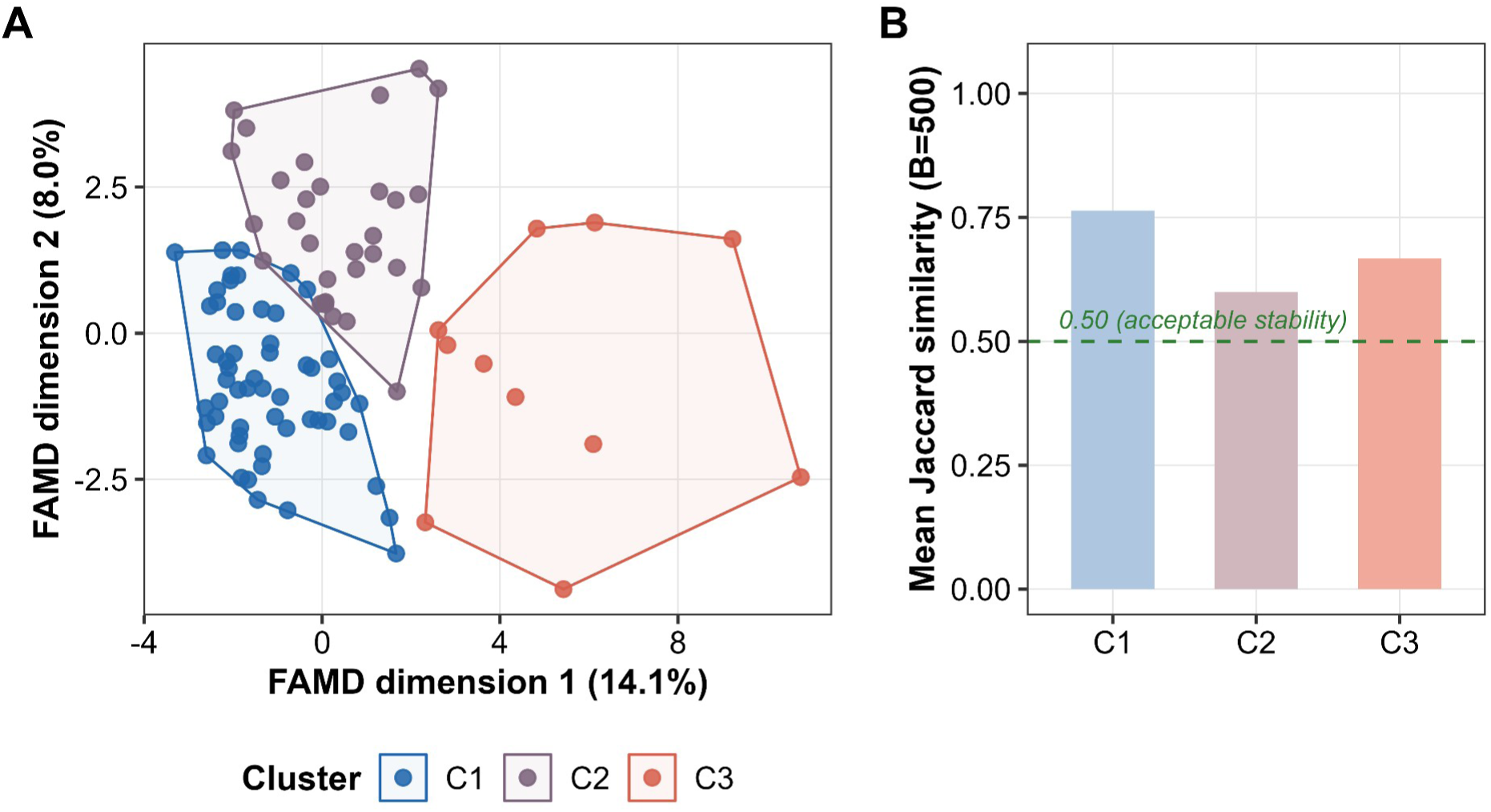
FAMD trajectory clusters and bootstrap stability. (A) Patient coordinates in the first two FAMD dimensions, coloured by trajectory cluster (C1, C2, C3); shaded regions show the convex hull of each cluster. (B) Mean bootstrap Jaccard similarity per cluster (B=500 resamples); a dashed line marks the pre-specified 0.50 acceptability threshold, met by all three clusters. Cohort: 98 patients (C1 n=56, C2 n=31, C3 n=11). FAMD Factor Analysis of Mixed Data.

### Cluster-Defining Burden Variables Follow a C1-to-C3 Gradient

We next evaluated if the clustering analysis could identify an increasing clinical burden across the C1-to-C3 gradient. To do so, four continuous variables shown (**Figure 2A-D**) were evaluated as the combination of age at diagnosis (younger age indicating higher burden), extra-intestinal manifestation (EIM) count, line-weighted therapy score and cumulative number of surgeries.

**Figure 2.**
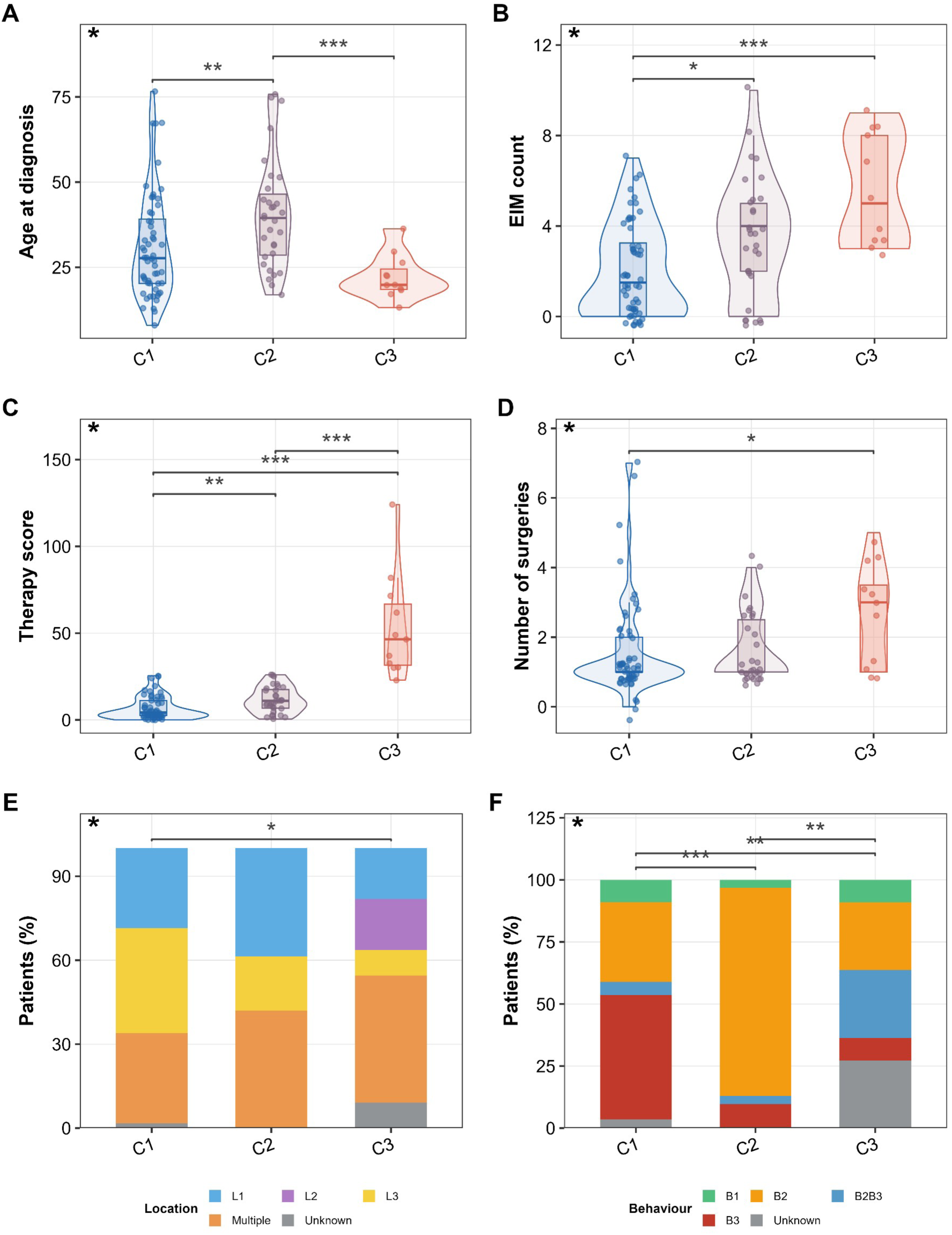
Disease-course phenotype and burden across trajectory clusters. (A) Age at diagnosis. (B) Extra-intestinal manifestation (EIM) count. (C) Therapy_Score. (D) Number of surgeries. (E) Montreal disease location. (F) Montreal behaviour. Panels A-D are shown as violin/box/jitter plots with Kruskal-Wallis tests; Panels E-F are shown as stacked percentage bars with chi-square tests; top-left asterisk marks the omnibus test’s Benjamini-Hochberg-adjusted P<0.05 (“at least one cluster differs”). Where the omnibus test was significant, brackets mark the specific cluster pair(s) that differ on post-hoc pairwise testing (Wilcoxon rank-sum for panels A-D, Fisher’s exact for panels E-F; Benjamini-Hochberg-adjusted within each variable’s three pairwise comparisons; * P<0.05, ** P<0.01, *** P<0.001); pairs not shown did not reach significance. EIM extra-intestinal manifestation; L1/L2/L3 Montreal disease location (ileal/colonic/ileocolonic); B1/B2/B3 Montreal behaviour (inflammatory/stricturing/penetrating); B2B3, coexisting stricturing and penetrating behaviour.

The clusters followed a clinically coherent gradient of disease-course burden (**Figure 2; Supplementary Table 2**), though not uniformly across every variable: C1 had older age at diagnosis, lower EIM burden and lower therapy burden, but a rate of prior surgery similar to C2 and C3 (see below); C2 showed intermediate burden; C3 was characterised by very young diagnosis age, high EIM burden, extensive advanced-therapy exposure and a greater cumulative surgery count (**Figure 2A-D**). Time to first biologic (log-rank P=0.326), time to Policlinico of Milan surgery (log-rank P=0.379) and any surgery by cluster (chi-square P=0.260) did not differ significantly across trajectory clusters; within this three-test family, all three Benjamini-Hochberg-adjusted P values converged to 0.379 **(Supplementary Figure 2**). Total surgical burden nonetheless differed significantly by cluster (Kruskal-Wallis P=0.024; median 1, 1 and 3 surgeries in C1, C2 and C3 respectively, **Table 1**), indicating that C3-cluster patients accumulate more surgeries over time without necessarily reaching their first surgery earlier or later than other clusters. The distinction between surgical burden and surgical timing is coherent with a picture in which C3-cluster patients undergo more aggressive medical/biologic escalation, which may delay individual surgical episodes without preventing the higher cumulative surgery count this subgroup ultimately accumulates (**Figure 2D**).

Comorbidity burden increased across the trajectory gradient too (**Supplementary Figure 3**). Autoimmune disease prevalence rose from 14.3% (C1) to 35.5% (C2) to 54.5% (C3), a difference that remained significant after Benjamini-Hochberg adjustment (P adj=0.009), and documented anxiety/depression increased in the same direction, from 11.3% to 23.3% to 36.4% in C1/C2/C3 respectively, though here the difference reduced its significance after adjustment (chi-square P=0.082, P adj=0.098; **Supplementary Figure 3A**). Three further comorbidities differed significantly by cluster after adjustment without following that same monotonic C1-to-C3 gradient: osteoporosis (C1 1.8%, C2 35.5%, C3 0.0%; P adj=0.003), hypothyroidism (C1 0.0%, C2 25.8%, C3 0.0%; P adj=0.005) and a history of infectious disease (C1 1.8%, C2 25.8%, C3 18.2%; P adj=0.005) were each most prevalent in C2 rather than C3 (**Supplementary Figure 3A**). Presenting gastrointestinal symptoms varied by cluster as well (**Supplementary Figure 3B**): fistulizing disease was, unexpectedly, most common in C1 (32.1%) rather than in C3 (9.1%) or C2 (3.2%) (P adj=0.018), mirroring the same paradoxical pattern seen for Montreal penetrating (B3) behaviour, while weight loss showed the expected gradient (8.9% in C1 to 36.4% in C3) without reaching significance after adjustment (P adj=0.063). Smoking status did not differ significantly by cluster (chi-square P=0.299; **Supplementary Figure 3C**), in keeping with its lack of association with baseline trajectory-membership screening reported below.

The three clusters resemble the Montreal disease classification [5] (**Figure 2E, F**), albeit B3 (penetrating) behaviour was, unexpectedly, most common in C1 (28/56) rather than in C3 (1/11), indicating that Montreal behaviour classification does not necessarily track the same disease-course gradient as cumulative treatment, surgical and comorbidity burden in this cohort.

### Cluster-Defining Advanced-Therapy Use Peaks in C3

Use rates differed significantly across clusters for most individual agents (chi-square; **Figure 3A, B; Supplementary Table 3**). Among conventional therapies, corticosteroid (P adj=0.034), mesalamine (P adj=0.049) and methotrexate (P adj<0.001) varied significantly across clusters, while azathioprine (P adj=0.079) exposure did not reach significance after adjustment (**Figure 3A**). Among advanced therapies, infliximab (P adj=0.004), vedolizumab (P adj=0.002), ustekinumab (P adj<0.001), upadacitinib (P adj<0.001) and risankizumab (P adj=0.004) all varied significantly, while adalimumab did not (P adj=0.124) despite being the most widely used agent overall (65.3%). C3-cluster patients showed the highest use rate for nearly every agent, most strikingly upadacitinib (36.4% vs 0-3.2% in C1/C2) and risankizumab (45.5% vs 3.2-10.7%), both reflecting this cluster’s more recent, treatment-refractory course **(Supplementary Figure 4A**).

**Figure 3.**
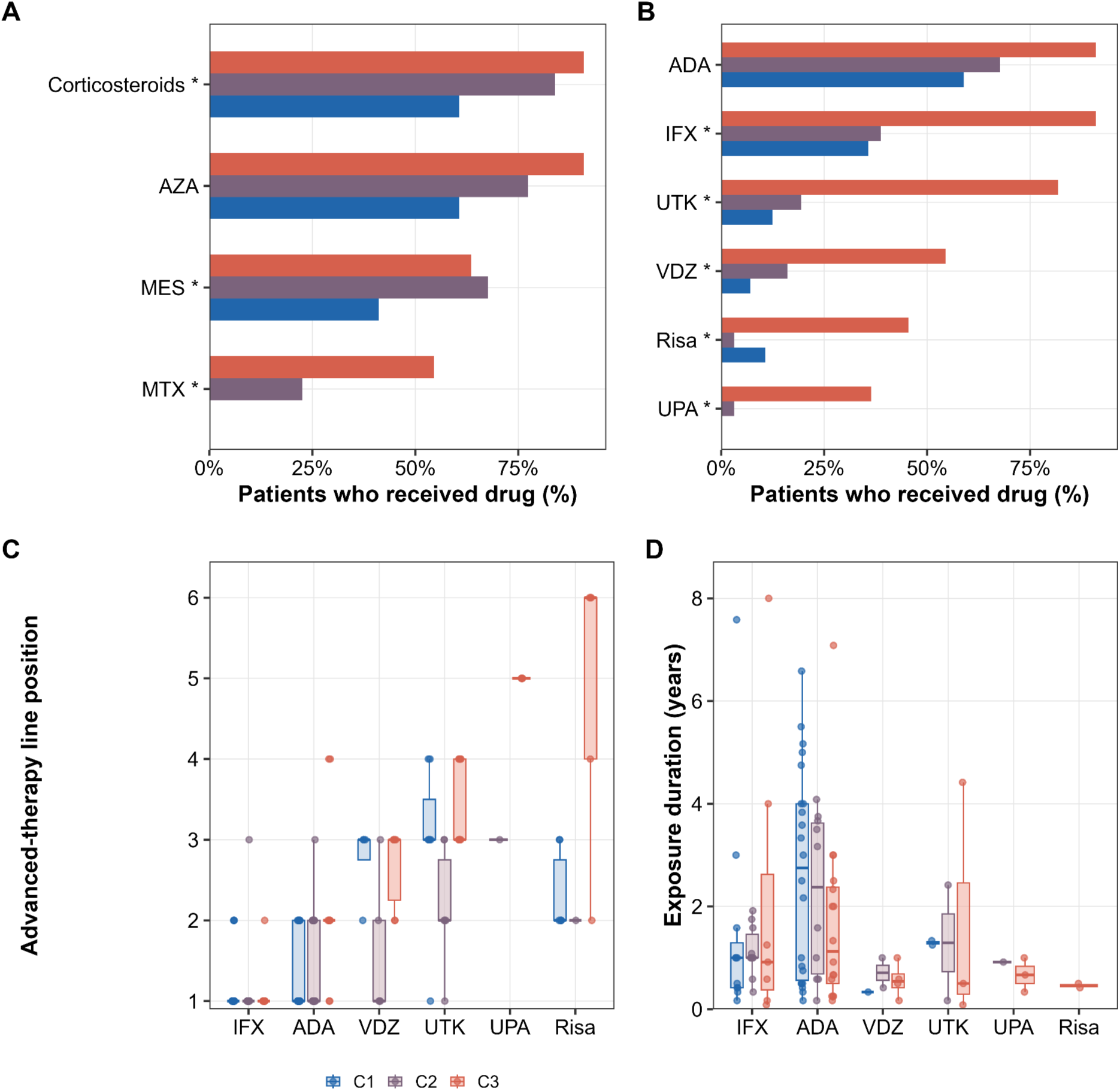
Advanced-therapy utilisation and sequencing. (A) Conventional therapy use rates by cluster (asterisk marks Benjamini-Hochberg-adjusted chi-square P<0.05). (B) Biologic and small-molecule use rates by cluster (asterisk marks Benjamini-Hochberg-adjusted chi-square P<0.05). (C) Advanced-therapy line position by drug and cluster. (D) Advanced-therapy exposure duration by drug and cluster. IFX, infliximab; ADA, adalimumab; VDZ, vedolizumab; UTK, ustekinumab; UPA, upadacitinib; Risa, risankizumab; Corticosteroidi, corticosteroids; MES, mesalamine; AZA, azathioprine; MTX, methotrexate.

Advanced-therapy line position also differed by cluster and agent (**Figure 3C; Supplementary Table 4)**: adalimumab was typically used as line 2 in C3 (median 2, IQR 2-2) versus line 1 in C1 and C2 (median 1, IQR 1-2), and risankizumab reached a later median line in C3 (line 6, IQR 4-6) than in C1 (line 2, IQR 2-2.8) (**Figure 3C**). Exposure duration varied by drug and cluster (**Figure 3D; Supplementary Table 5**), with generally shorter durations in C3 (e.g., adalimumab median 1.13 years (IQR 0.5-2.37) in C3 versus 2.75 years (IQR 0.56-4) in C1), in keeping with more frequent treatment switching in this cluster.

### Treatment Era Differs Significantly by Cluster

The advanced-therapy armamentarium available to CD patients expanded substantially over the study period, and loss of response to biologic therapy is a well-recognised but variable feature of Crohn’s disease management [2–4]. To test whether clusters differed in when episodes of therapeutic failure began, relative to each drug’s own approval, we analysed the treatment era. Each episode was classified relative to that specific drug’s own approval date as pre-approval, early (0-4 years post-approval), established (5-9 years) or mature (≥10 years) (**Supplementary Figure 4B**). Because a single patient can contribute multiple episodes, treatment era distribution was compared as a Mature-vs-not-yet-mature indicator using a GEE clustered by patient; it differed significantly by cluster (P=0.019; **Figure 4A; Supplementary Table 6**): C3-cluster episodes were concentrated in the early post-approval era (35.8% early vs 9.9% in C1 and 12.2% in C2), while C1 and C2 episodes were predominantly in the mature era (62.0% and 59.2% respectively, vs 30.2% in C3). That skew was not confined to the most recently approved agents: only 8 of C3’s 20 early/pre-approval episodes were on upadacitinib or risankizumab, while the remaining 12 were early-era episodes of adalimumab, ustekinumab, vedolizumab and infliximab, agents with an established mature-era track record elsewhere in this cohort (**Supplementary Table 7**). Such a pattern is consistent with C3’s high clinical burden driving earlier and broader use of recently approved advanced therapy, including agents whose own optimal dosing and administration were possibly still being defined.

**Figure 4.**
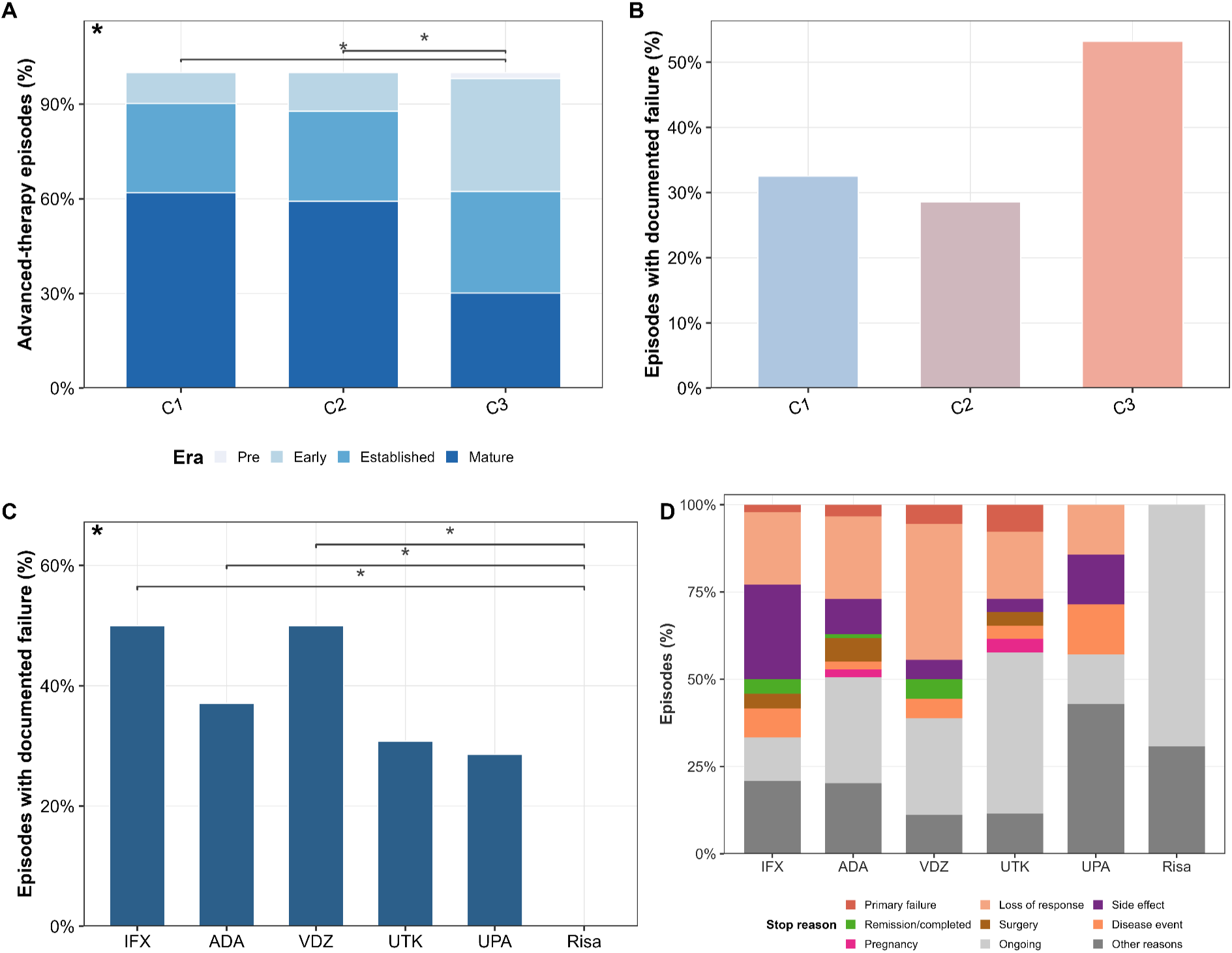
Durability and documented failure of advanced therapy. (A) Treatment era distribution by cluster (top-left asterisk marks P<0.05 from a GEE clustered by patient). (B) Episodes with documented failure (primary failure, loss of response or side effect) by cluster (episodes are correlated within patient; a GEE clustered by patient gave P=0.085, so no asterisk is shown). (C) Episodes with documented failure by individual drug, pooled across clusters (top-left asterisk marks the omnibus chi-square P<0.05). Where the omnibus test was significant, brackets mark the specific group pair(s) that differ on post-hoc pairwise testing (GEE-based contrasts for panel A, Fisher’s exact for panel C; Benjamini-Hochberg-adjusted within each panel’s comparisons; * P<0.05, ** P<0.01, *** P<0.001); pairs not shown did not reach significance. (D) Reason for discontinuation by advanced-therapy agent, all episodes (still-active episodes grouped as Ongoing, unclassifiable stop text as other reasons). IFX, infliximab; ADA, adalimumab; VDZ, vedolizumab; UTK, ustekinumab; UPA, upadacitinib; Risa, risankizumab.

Episodes with documented failure (primary failure, loss of response or side effect) were more frequent in C3 (53.2%, 33/62) than in C1 (32.5%, 27/83) or C2 (28.6%, 16/56; **Figure 4B; Supplementary Table 8**). A single patient can contribute several episodes, though, so a GEE clustered by patient did not reach conventional significance (P=0.085). Episodes concentrated in each drug’s own early-use window may help explain C3’s higher documented-failure rate, reflecting both a difficult-to-treat patient population and treatment given before that drug’s posology had been fully optimised.

Documented failure also differed significantly by individual drug, pooled across clusters (chi-square P=0.022; **Figure 4C; Supplementary Table 9**): infliximab (50.0%, 24/48) and vedolizumab (50.0%, 9/18) showed the highest failure rates, adalimumab (37.1%, 33/89) and ustekinumab (30.8%, 8/26) intermediate rates, and risankizumab showed 0 of 13 documented failures. Such an apparently favourable risankizumab profile possibly reflects its recent approval (2022) and correspondingly a shorter follow-up rather than superior effectiveness (**Supplementary Figure 4C**).

Reason for treatment discontinuation varied substantially by agent (**Figure 4D; Supplementary Table 10**): infliximab and vedolizumab showed the highest combined loss-of-response/side-effect burden (47.9% and 44.4% of episodes respectively), while risankizumab showed no documented failure of any kind, with the large majority of episodes (69.2%, 9/13) still ongoing at data cut-off, in keeping with its recent approval and correspondingly short follow-up.

### Diagnosis-Time Variables Are Associated With the C3 Trajectory

As we aimed at identifying candidate predictors associated with trajectory membership, we performed a univariate screening analysis of diagnosis-available variables. Six variables were identified as significant after Benjamini-Hochberg adjustment (**Figure 5A; Table 2**). Among them Montreal behaviour (P adj<0.001, Cramér’s V=0.478) and associated comorbidities (P adj<0.001, V=0.414) were the strongest correlates, ahead of EIM count (P adj<0.001; per-EIM odds ratio 1.55, 95% CI 1.21-2.08) and age at diagnosis (P adj=0.001; per-year odds ratio 0.93, 95% CI 0.85-0.98). Montreal disease location (P adj=0.004) and Montreal age class (P adj=0.016) were also identified as significant. Sex (P adj=0.224) and smoking status (P adj=0.276) were not significantly associated with cluster membership.

**Figure 5.**
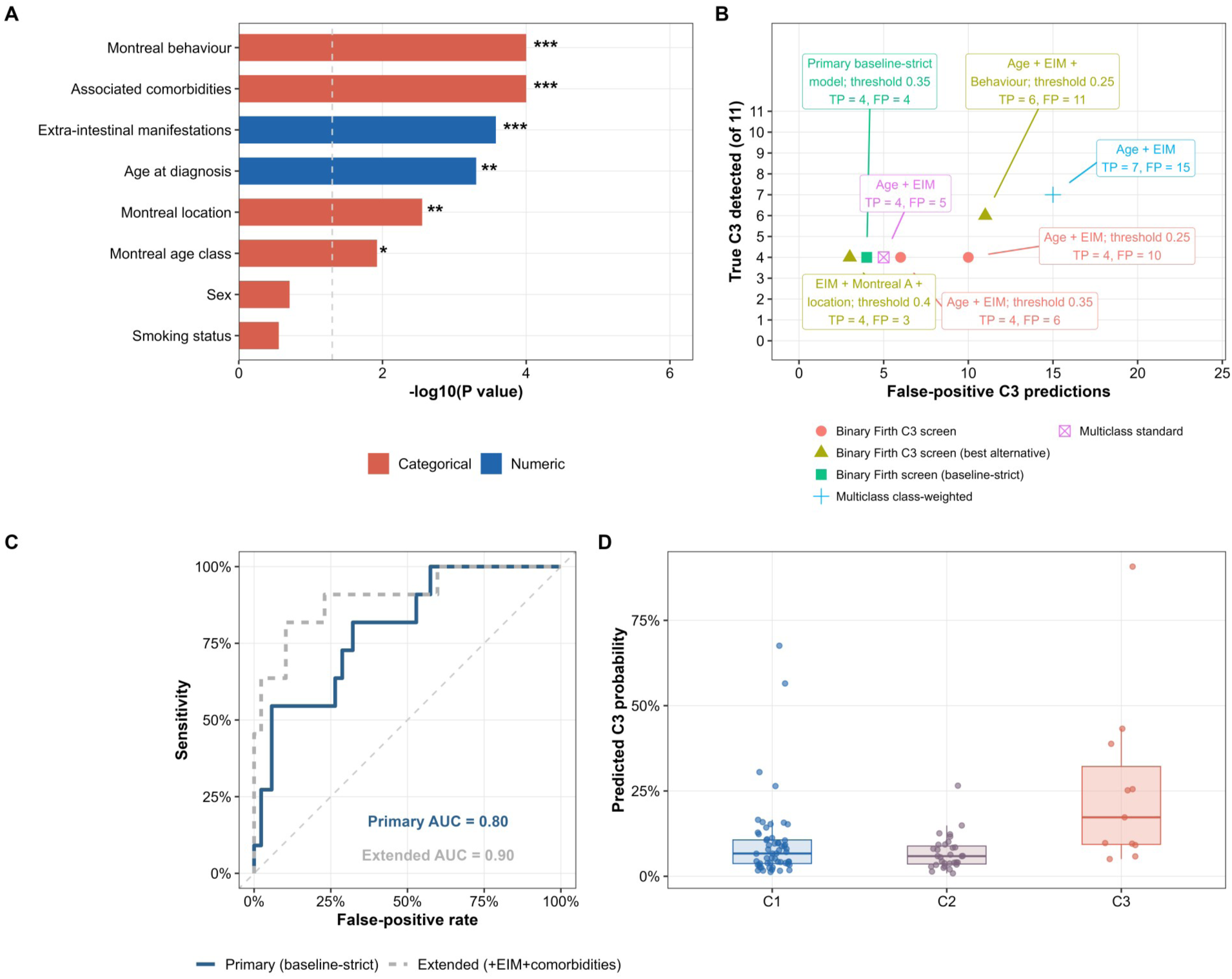
Baseline prediction of disease-course trajectory cluster. (A) Univariate candidate-predictor screening (bar length = -log10 raw P value; asterisk marks Benjamini-Hochberg-adjusted P<0.05, **<0.01, ***<0.001). (B) Internal C3-detection trade-off across candidate screening approaches (true C3 detected vs false-positive C3 predictions). (C) ROC curves for the primary model (age at diagnosis, disease site, Behaviour) and the extended sensitivity model (adding EIM count and associated comorbidities), both leave-one-out cross-validated ridge logistic regressions. (D) Predicted C3 probability by cluster from the primary model. EIM extra-intestinal manifestation.

We compared the internal C3-detection trade-off across several candidate screening approaches, binary Firth logistic models combining age at diagnosis, EIM count and, variably, Montreal_A, disease site or Behaviour (**Figure 5B; Supplementary Table 11**); adding Behaviour to the age-and-EIM screen improved detection overall. The best-precision alternative (EIM + Montreal_A + disease location, threshold 0.40) detected 4/11 C3 patients with only 3 false positives, and the best-sensitivity alternative (age + EIM + Behaviour, threshold 0.25) detected 6/11 C3 patients (54.5%) at a cost of 11 false positives. Class-weighted multiclass and random-forest approaches using only age, EIM and (for random forest) location and sex performed less favourably (4-7 of 11 C3 patients detected at 4-15 false positives).

Of the diagnosis-available candidates, only Age_at_Diagnosis, Disease_Site and Behaviour reached Benjamini-Hochberg-adjusted significance in univariate screening without raising the timing or redundancy concerns discussed above (**Table 2**). This combination was fixed before any multivariable model was fitted, and defines the primary predictive model. A leave-one-out cross-validated ridge logistic regression on these three variables (**Table 3**) reached an AUC of 0.80 (95% CI 0.67-0.93) and a Brier score of 0.084 (**Figure 5C**), with good calibration (intercept 0.08, slope 1.00). It captured 3 of the top-5 and 5 of the top-10 highest-risk patients as true C3 cases; the mean predicted C3 probability was 0.255 among observed C3 patients versus 0.087 among non-C3 patients (**Figure 5D; Supplementary Table 12**). Adding EIM count and associated comorbidities in a second, extended model raised discrimination to an AUC of 0.90 (95% CI 0.79-1.00; **Figure 5C**). Because the timing of both variables relative to diagnosis could not be confirmed, we report this only as a sensitivity analysis, one that suggests a meaningful share of the extra signal in the less restricted model comes from disease-course information that only accrues after diagnosis.

**Table 3.** Baseline predictive model performance: primary (diagnosis-available) model and extended sensitivity model.

| Model | Variables | N | C3 N | AUC | Brier | Top-5 C3 | Top-10 C3 | Status |
| --- | --- | --- | --- | --- | --- | --- | --- | --- |

|  |  |  |  | (95%<br>CI) |  | captured | captured |  |
| --- | --- | --- | --- | --- | --- | --- | --- | --- |
| Primary model | Age at diagnosis +<br>disease site + Behaviour | 98 | 11 | 0.80<br>(0.67<br>-<br>0.93) | 0.084 | 3 | 5 | LOOCV ridge logistic<br>model; calibration<br>intercept 0.08, slope<br>1.00 |
| Extended model<br>(sensitivity<br>analysis) | Age at diagnosis +<br>disease site + Behaviour<br>+ EIM count + associated<br>comorbidities | 98 | 11 | 0.90<br>(0.79<br>-<br>1.00) | 0.059 | 5 | 7 | LOOCV ridge logistic<br>model; calibration<br>intercept 0.46, slope<br>1.19; not the primary<br>claim - EIM and<br>comorbidity timing<br>relative to diagnosis is<br>not confirmed |

## Discussion

Crohn’s disease is a heterogeneous condition whose clinical course varies widely even among patients who appear similar at diagnosis, a variability that conventional, single-time-point classification systems are not designed to capture. Here we identified three CD-course trajectory phenotypes that were reproducibly recovered under bootstrap resampling (Jaccard 0.60-0.76 across clusters), revealing a gradient from lower-burden disease to a small but highly refractory C3 subgroup. Their internal geometric separation was more modest (average silhouette width 0.29), which is more consistent with a disease-course burden behaving as a continuum rather than a fixed three-way partition. Differently from prior unsupervised-clustering work in inflammatory bowel disease [7,8], we analysed the full disease-course trajectory of an actively treated cohort, and explicitly tested whether Montreal-classification-era variables can anticipate the resulting phenotype, a two-step design that, to our knowledge, has not previously been applied specifically to CD disease-course burden.

Our trajectory-based approach differs from the Montreal and Paris classifications. The Montreal classification stratifies Crohn’s disease patients by age at diagnosis, disease location and behaviour, and remains the reference standard for phenotypic description in clinical practice and clinical trials [5]; a paediatric adaptation, the Paris classification, extends this framework with growth-impairment and age-of-onset categories relevant to childhood-onset disease [6]. Those systems assign a category at a single time point, typically at or near diagnosis, whereas the clusters identified here are defined by cumulative treatment, surgical and comorbidity history accrued over a median disease duration of nearly 18 years. In this context the C3 trajectory could not have been identified from Montreal criteria alone: Montreal behaviour at diagnosis disagreed with cluster membership for a sizeable subset of patients, because behaviour, location and age class describe disease presentation rather than the treatment-intensive course that later unfolds. The distinction matters clinically as much as methodologically: treating a single diagnosis-time snapshot as a proxy for the whole disease course risks missing exactly the kind of divergence this cohort shows, since two patients who look alike on Montreal criteria at diagnosis can accumulate very different burdens of treatment escalation, surgery and comorbidity, and only a framework that follows the course itself, rather than inferring it from baseline features, can tell them apart. In CD patients, disease behaviour is known to evolve over time rather than remain fixed at diagnosis: in the Vienna-classification cohort, nearly half of patients changed their behaviour category (most commonly from inflammatory to stricturing or penetrating disease) over ten years of follow-up [24]. That natural evolution in behaviour classification over time may help explain why B3 (penetrating) behaviour was, in this cohort, more common in the lower-burden C1 trajectory than in C3. Patients whose penetrating complication was addressed by early definitive surgery may show a more indolent subsequent medical course. At the same time, progressive multidrug escalation, as captured by the C3 trajectory regardless of initial behaviour classification, may reflect an independent, cumulative dimension of disease severity that Montreal behaviour recorded at a single time point does not fully capture. Population-based natural-history data indicate that roughly half of CD patients develop an intestinal complication within 20 years and half require surgery within 10 years [25], with younger age at diagnosis, extensive small-bowel disease, perianal involvement, early stricturing/penetrating behaviour and corticosteroid requirement at first flare consistently identified as predictors of a more complicated course [26,27]. However, baseline clinical parameters alone have previously been shown insufficient to reliably predict a disabling course [28], and registry-based multivariable prediction models for CD complications remain a relatively recent and still-evolving approach [29]. The data-driven trajectory phenotypes identified in our study are consistent with this literature: rather than assuming a single trajectory implied by baseline classification alone, unsupervised clustering on the full disease-course history identified a coherent gradient culminating in the small, high-burden C3 subgroup described above, reinforcing the rationale for a two-step framework that first characterises realised disease-course trajectories and only then asks whether diagnosis-available variables can anticipate them.

Age at diagnosis and EIM burden were among the strongest baseline correlates of trajectory membership in the C3 cohort, consistent with prior reports identifying young age at diagnosis and extraintestinal manifestations among the most reproducible predictors of a more aggressive CD course [26,27]. As in prior work [28], no single baseline model reliably identified the smallest, most severe subgroup without a material false-positive cost in internal cross-validation, even after adding the strongest available categorical predictor (Behaviour); this underscores that baseline clinical variables offer exploratory discrimination rather than definitive early classification, at least in cohorts of this size. Our estimate is lower than that of another recently published prediction model for complicated CD: a population-based study combining clinical, serological and genetic predictors in paediatric-onset Crohn’s disease reported an AUC of 0.84 (95% CI 0.78-0.90) for a 5-year complicated course [30], somewhat higher than our primary model despite drawing on a broader predictor set. Our model uses only three routinely recorded clinical variables, with no laboratory or genetic testing, in a different population (adult-onset, single-centre); the comparison is therefore limited, but it suggests that a small set of already-collected clinical variables can approach the discriminative performance of considerably more elaborate signatures. These findings can also be read against the recently proposed “difficult-to-treat” inflammatory bowel disease framework: an international consensus formally defined this phenotype as persistent disease activity despite sequential exposure to multiple therapeutic classes [10], and a subsequent real-world cohort study characterised its prevalence, clinical features and outcomes [11]. The C3 trajectory identified here, marked by early disease onset, a heavy extra-intestinal manifestation burden and resistance to treatment, shares clinical features that overlap with that difficult-to-treat phenotype, rather than being demonstrably the same population: C3 was derived by unsupervised, data-driven clustering, not validated against the sequential multi-class-exposure definition the consensus criteria actually specify, so the overlap remains a hypothesis that requires direct external validation against those criteria.

Autoimmune disease and anxiety/depression were both most prevalent in the C3 phenotype. Indeed anxiety and depression are increasingly recognised as common comorbidities of inflammatory bowel disease, possibly through microbiota-gut-brain axis signalling [31]; the autoimmune-disease gradient remained significant after multiplicity adjustment in this cohort, and both patterns support collecting mental-health and multisystem comorbidity outcomes alongside gastrointestinal measures in future prospective work on this phenotype. This multisystem burden pattern points to a concrete, testable use case: flagging patients who screen as C3-like at diagnosis (**Figure 5B**) for earlier multidisciplinary referral (rheumatology, dermatology, ophthalmology, mental-health support) alongside gastroenterology follow-up. This is a hypothesis, not a tested finding, and would need prospective evaluation before anyone could claim it improves outcomes.

We also evaluated how therapies may impact on the trajectory phenotype definition. C3 patients receive therapies at later lines and have more documented primary failure, loss of response or side effect. Such variable durability echoes a broader literature on loss of response to biologic therapy in Crohn’s disease: a meta-analysis pooling 55 studies estimated anti-TNF loss of response at 33% (95% CI 29-38) at a median of one year of follow-up, with individual study estimates ranging from 8% to 71% [2]; comparable variability extends to newer, non-anti-TNF agents, with one multicentre cohort reporting a 36.7% (95% CI 26.6-49.2%) probability of composite loss of response to vedolizumab by week 52 in CD [3]. That variability is not confined to loss of response on a single agent: a population-based study across Catalonia documented substantial geographic variation in which pharmacological therapies were prescribed for Crohn’s disease and in the outcomes achieved [4]. However, one has to take treatment-era into account to fully compare documented failure or loss of response across drugs and clusters, and in two distinct ways. First, C3’s numerically higher documented-failure rate (not statistically confirmed once multiple episodes per patient are accounted for, see Results) may partly reflect the exploratory use of new available drugs in this type of multifailure patients: its upadacitinib and risankizumab episodes, together with 12 of its 20 early/pre-approval episodes on adalimumab, ustekinumab, vedolizumab and infliximab (**Supplementary Table 7**), began while that specific drug’s own dosing and administration practice was still maturing. Post-marketing dosing changes to biological product labels are reported a median of four years after approval (range 1-7 years) [32]. Second, and working in the opposite direction, risankizumab, the most recently approved biologic in this cohort, showed 0 of 13 documented failures (9 episodes still ongoing at data cut-off, 4 stopped for other non-failure reasons; **Figure 4C, D**; **Supplementary Table 9**, 10); its apparently low failure rate more likely reflects limited follow-up time than superior effectiveness, and should not be compared directly with agents used for longer in this cohort. With the data available here, both mechanisms may contribute to some degree, and future work with larger, prospectively followed cohorts would help clarify how much of C3’s elevated documented-failure rate reflects genuine treatment-refractoriness versus these era-related confounds.

In summary, this study benefits from several methodological strengths that support confidence in its findings: a two-step analytic design that separates data-driven phenotyping from diagnosis-available prediction, cluster robustness confirmed through two independent validation methods, and rigorous statistical handling of rare-event bias and multiple testing. These strengths, however, must be weighed against important limitations. The clustering solution itself has only moderate internal validity by conventional benchmarks: the three retained FAMD dimensions captured just 28.2% of total variance in the active variable set, and the average silhouette width of 0.29 falls within the range conventionally interpreted as weak or artificial rather than strong natural structure, a pattern also reported in comparable clinical cohort clustering studies on mixed-type data (e.g. a silhouette width of 0.29 in a FAMD-based COVID-19 phenotyping study of 547 patients [33]), consistent with disease severity behaving as a continuum rather than naturally discrete categories; the three trajectories should therefore be read as one specific, low-dimensional summary of disease-course heterogeneity rather than an exhaustive or uniquely correct partition of it. The retrospective, single-centre design, combined with a small C3 cluster (n=11), limits the generalisability and statistical power of class-specific inferences. Because the cohort was ascertained through a surgical/IBD referral pathway at the IRCCS Policlinico of Milan, the high proportion of patients with recorded surgery likely reflects referral bias rather than the true surgical burden of a general CD population, and findings related to surgery and comorbidity should be interpreted accordingly. Advanced-therapy stop-reason categorisation was similarly limited by the completeness of clinical documentation at the time of discontinuation. Disease activity itself was captured only through treatment, surgical and comorbidity proxies: objective inflammatory markers such as C-reactive protein and faecal calprotectin, and endoscopic disease activity data, were not available in this dataset and so could not be incorporated into either the trajectory phenotyping or the baseline prediction model. Documented failure was ascertained retrospectively from clinical charts as a single composite (primary failure, loss of response or side effect), without a minimum follow-up window or a formal time-to-failure analysis. Episodes still ongoing at data cut-off were counted as non-failure, a form of right-censoring that likely understates failure for more recently approved agents such as risankizumab. Taken together, these limitations underscore the need for external validation in an independent, non-surgically-referred cohort: such validation would clarify whether the C3 trajectory and its associated clinical correlates represent a real disease-course phenotype rather than a referral-driven artefact, would test whether the baseline model’s promising discriminative performance (AUC=0.80 for the primary model, 0.90 for the extended sensitivity model) and probability thresholds generalise beyond internal cross-validation, and would help distinguish treatment-era effects from true phenotype-driven differences in outcome.

### Conclusions

Data-driven mixed-data clustering identified stable CD-course trajectory phenotypes. Age at diagnosis, disease location and Montreal behaviour, the diagnosis-available variables retained after univariate screening, showed exploratory discrimination for the high-burden C3 trajectory. EIM burden and comorbidity added further discrimination, but only in a sensitivity analysis whose timing relative to diagnosis we could not confirm. External validation and careful timing verification are required before clinical implementation. These results illustrate the value of disease-course phenotyping as a discovery tool for patterns that baseline-only analyses would miss.

## Author Contributions

FC, FF conceived the study. FF, FC, BC designed the study. FF, FC designed and supervised the experiments and analyses. CA, LP, AA generated the databases, DN, LBo, LBa, SC, MP, BO enrolled and treated patients. BC, FF wrote the draft. DN, CA, FC, FF edited the final version. All authors reviewed and critically edited the manuscript. All authors contributed to the article and approved the submitted version.

## Data Availability

The authors confirm that the data supporting the findings of this study are available within the article and its supplementary materials. The de-identified dataset and the full analysis pipeline (R scripts) are additionally available at https://github.com/bruneja/acdc-crohn-trajectory-phenotyping. The shared dataset is the same workbook used for the analyses reported here, de-identified by generalising exact dates to the first of January of the recorded year and removing free-text clinical notes.

## Declaration of Generative AI and AI-Assisted Technologies in the Writing Process

During the preparation of this manuscript, the authors used Claude (Anthropic), a large language model-based AI assistant, to support development of the reproducible analysis pipeline, and editing of manuscript text. After using this tool, the authors reviewed, verified and edited all AI-assisted content, and take full responsibility for the accuracy, integrity and scientific content of this publication.

## SUPPLEMENTARY MATERIAL

### Supplementary Figures

**Supplementary Figure 1.**
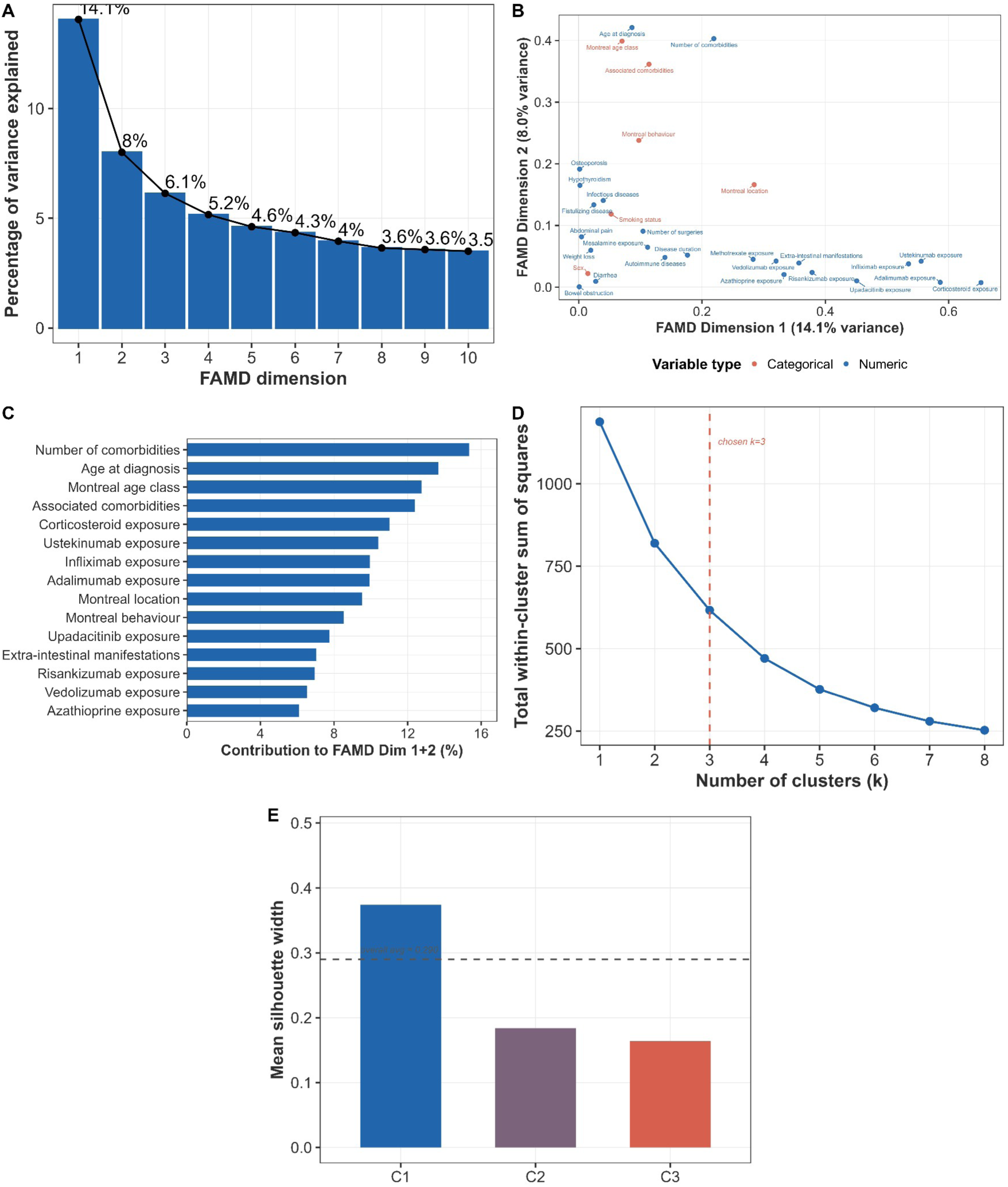
FAMD trajectory clustering methodology. (A) Scree plot of variance explained by each FAMD dimension. (B) FAMD variable map for active variables on Dimensions 1-2. (C) Variable contributions to FAMD Dimensions 1-2. (D) Within-cluster sum-of-squares elbow plot across candidate cluster counts. (E) Per-cluster silhouette width across candidate cluster counts (k).

**Supplementary Figure 2.**
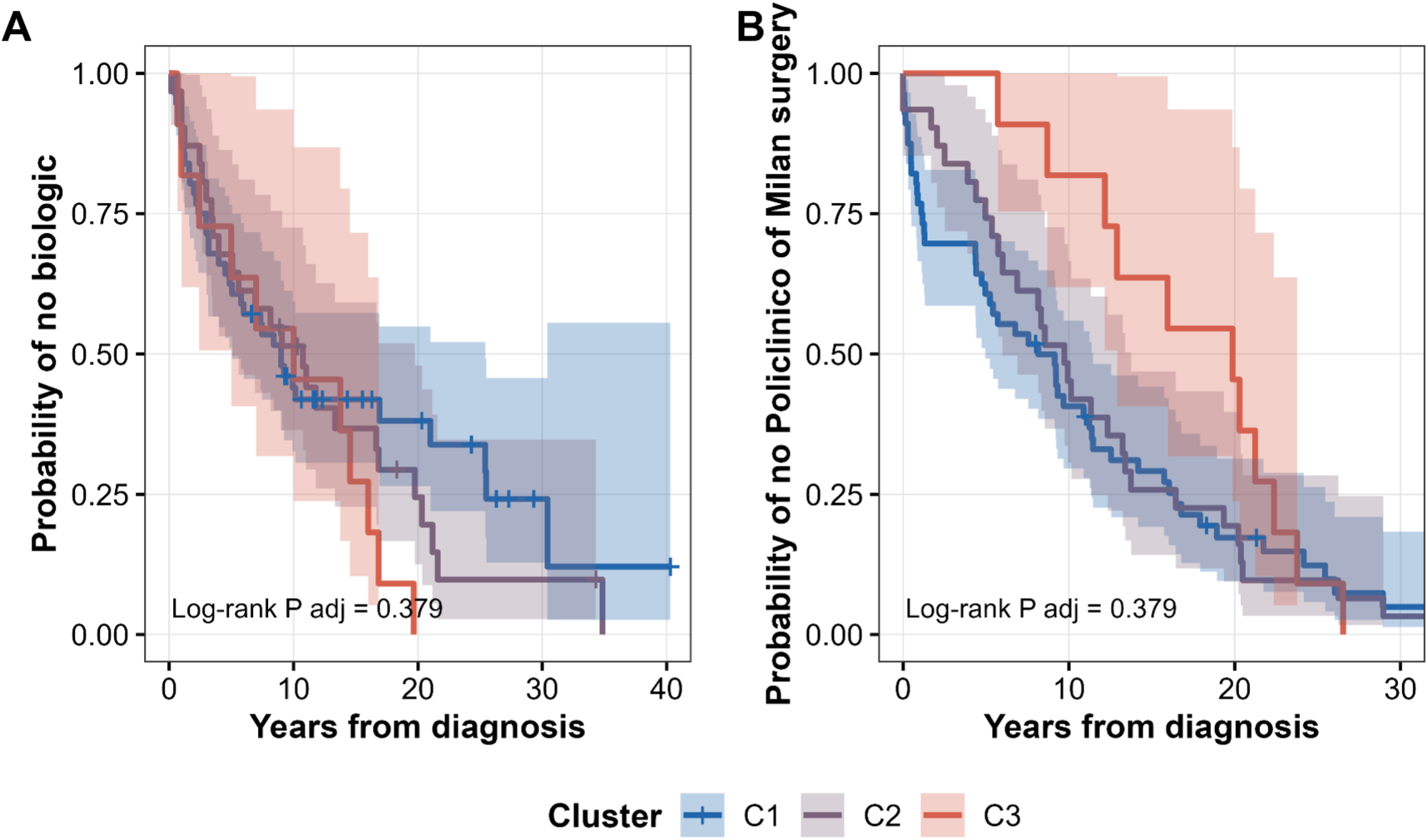
Time to first biologic and time to surgery by cluster. (A) Kaplan-Meier time-to-first-biologic curves by cluster. (B) Kaplan-Meier time-to-Policlinico-di-Milano-surgery curves by cluster. Neither comparison reached significance after Benjamini-Hochberg adjustment (time to first biologic: log-rank P=0.326, P adj=0.379; time to surgery: P adj=0.379).

**Supplementary Figure 3.**
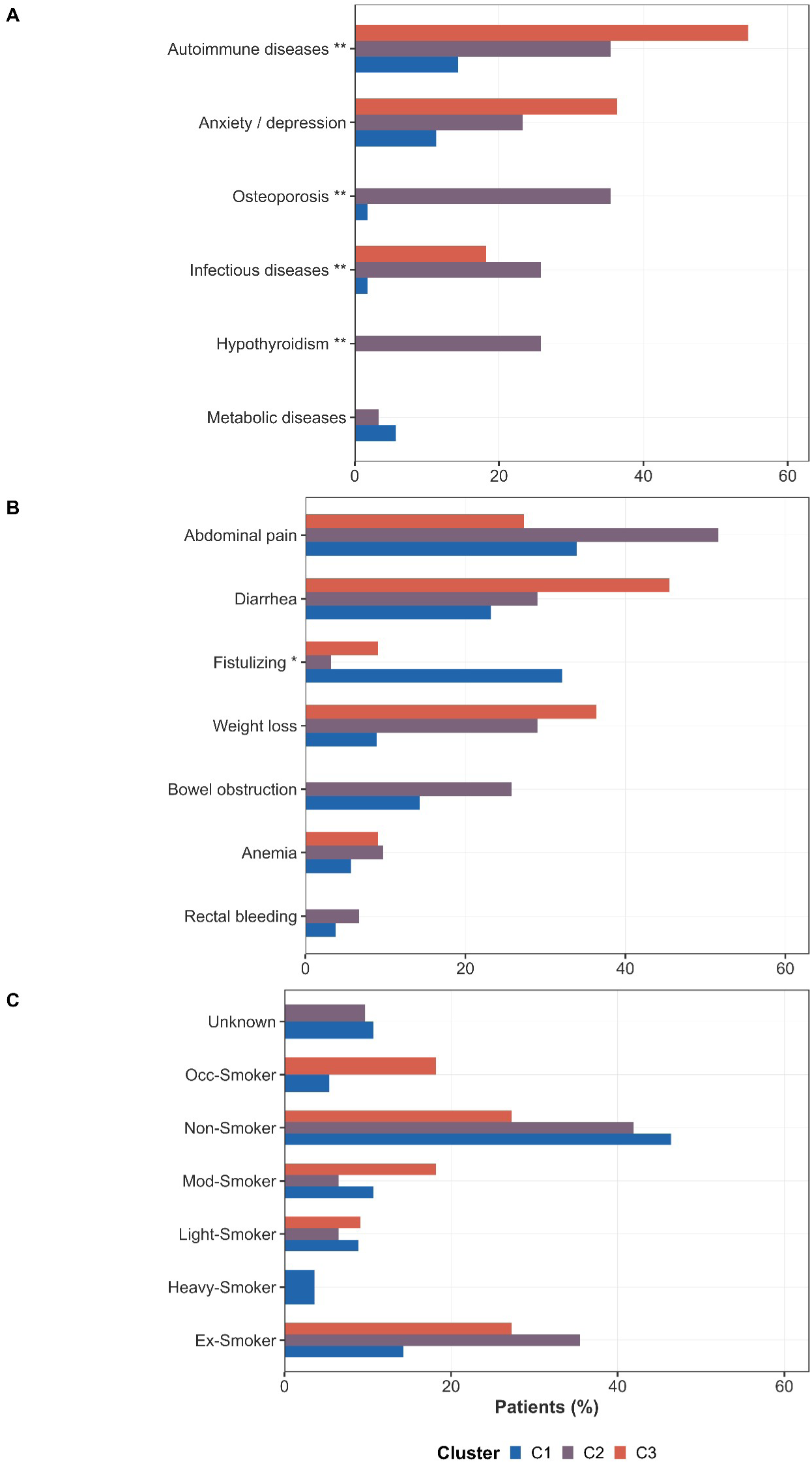
Comorbidities, smoking status and presenting symptoms by cluster. (A) Comorbidity prevalence by cluster. (B) Presenting gastrointestinal symptoms by cluster. (C) Smoking status by cluster.

**Supplementary Figure 4.**
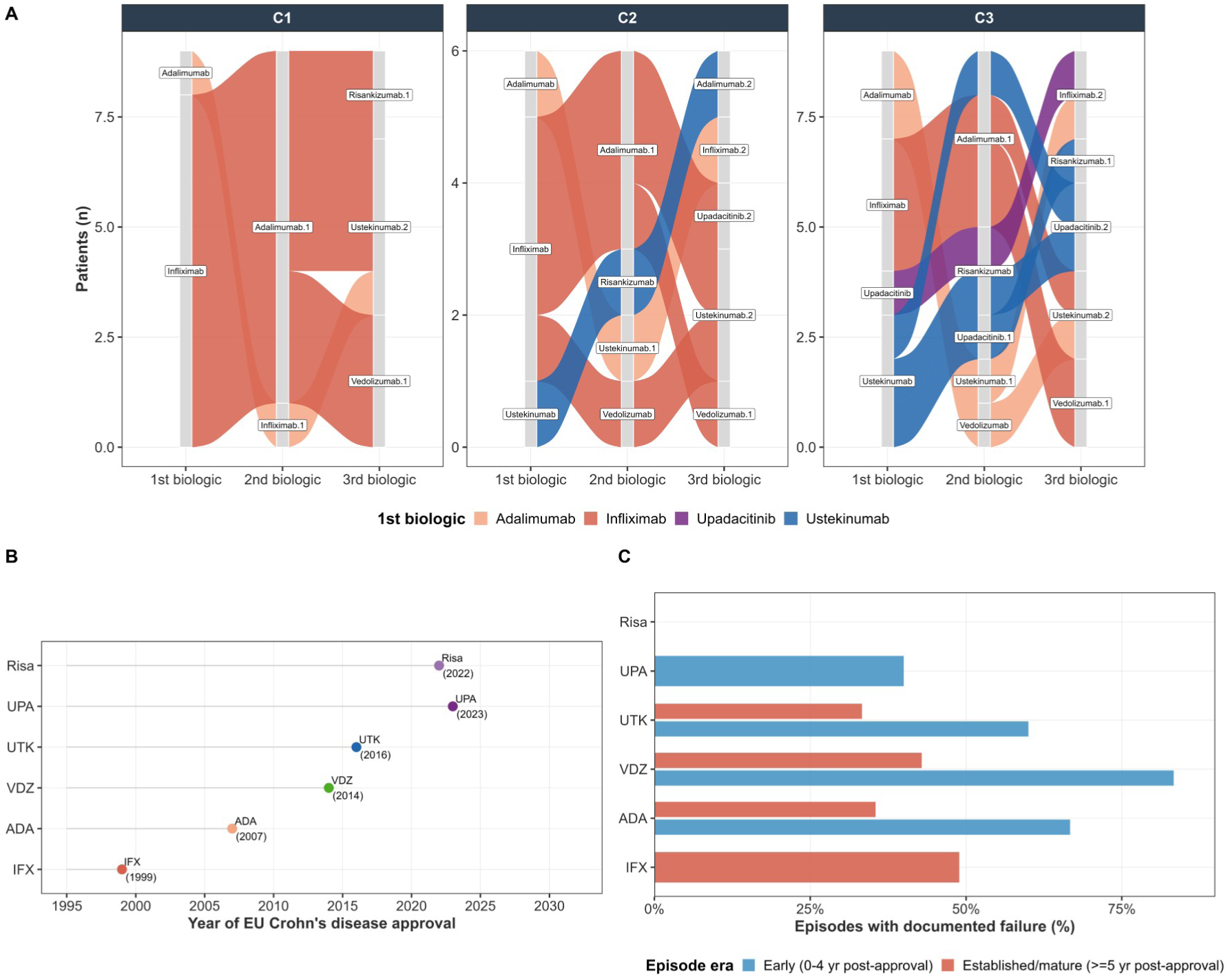
Biologic sequencing and treatment-era context. (A) Alluvial diagram of biologic sequencing by cluster. (B) EU approval timeline for each advanced-therapy agent. (C) Treatment-era-stratified failure rates.

### Supplementary Tables

**Supplementary Table 1.** Cluster size and bootstrap stability.

| Cluster | N | Bootstrap Jaccard |
| --- | --- | --- |
| C1 | 5<br>6 | 0.76 |
| C2 | 3<br>1 | 0.60 |
| C3 | 1<br>1 | 0.67 |

*N* is the number of patients per trajectory cluster (total $N=98$ ). Bootstrap Jaccard is the mean per-cluster Jaccard similarity across $B=500$ bootstrap resamples (*kmeansCBI*); values above 0.50, the dissolution threshold of Hennig (2007), were considered acceptable.

**Supplementary Table 2.** Statistical tests underlying Figure 2 (disease-course burden by cluster).

| Variable (Figure 2 panel) | P value | P adj (BH) | Test |
| --- | --- | --- | --- |
| Age at diagnosis | <0.001 | 0.001 | Kruskal-Wallis |
| EIM count | <0.001 | <0.001 | Kruskal-Wallis |
| Therapy_Score | <0.001 | <0.001 | Kruskal-Wallis |
| Number of surgeries | 0.024 | 0.024 | Kruskal-Wallis |
| Montreal disease location | 0.003 | 0.004 | Chi-square simulated |
| Montreal behaviour | <0.001 | <0.001 | Chi-square simulated |

**Supplementary Table 3.** Per-drug use rates by cluster.

| Drug | Type | Overall n (%) | C1 n (%) | C2 n (%) | C3 n (%) | P adj (chi-sq, BH) |
| --- | --- | --- | --- | --- | --- | --- |
| IFX | Biologic | 42 (42.9%) | 20 (35.7%) | 12 (38.7%) | 10 (90.9%) | 0.0043 |
| ADA | Biologic | 64 (65.3%) | 33 (58.9%) | 21 (67.7%) | 10 (90.9%) | 0.1235 |
| VDZ | Biologic | 15 (15.3%) | 4 (7.1%) | 5 (16.1%) | 6 (54.5%) | 0.0018 |
| UTK | Biologic | 22 (22.4%) | 7 (12.5%) | 6 (19.4%) | 9 (81.8%) | 5e-04 |
| UPA | Small molecule | 5 (5.1%) | 0 (0%) | 1 (3.2%) | 4 (36.4%) | 5e-04 |
| Risa | Biologic | 12 (12.2%) | 6 (10.7%) | 1 (3.2%) | 5 (45.5%) | 0.0043 |
| Corticosteroids | Conventional | 70 (71.4%) | 34 (60.7%) | 26 (83.9%) | 10 (90.9%) | 0.0344 |
| MES | Conventional | 51 (52%) | 23 (41.1%) | 21 (67.7%) | 7 (63.6%) | 0.0492 |
| AZA | Conventional | 68 (69.4%) | 34 (60.7%) | 24 (77.4%) | 10 (90.9%) | 0.0793 |
| MTX | Conventional | 13 (13.3%) | 0 (0%) | 7 (22.6%) | 6 (54.5%) | 7e-04 |

Type: Biologic, small molecule or conventional therapy. P adj is Benjamini-Hochberg-adjusted chi-square P (simulated) across the three clusters.

**Supplementary Table 4.**
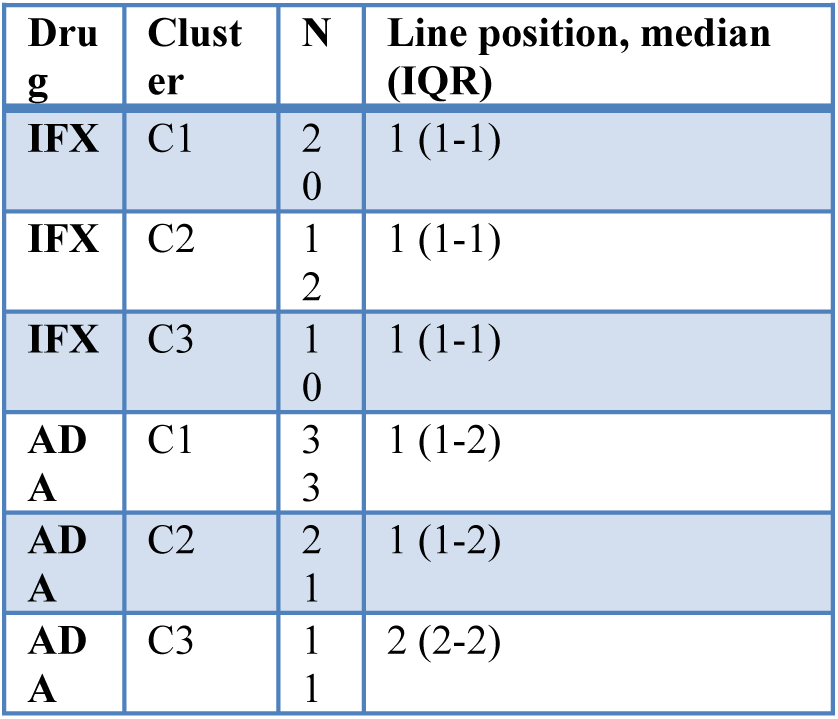
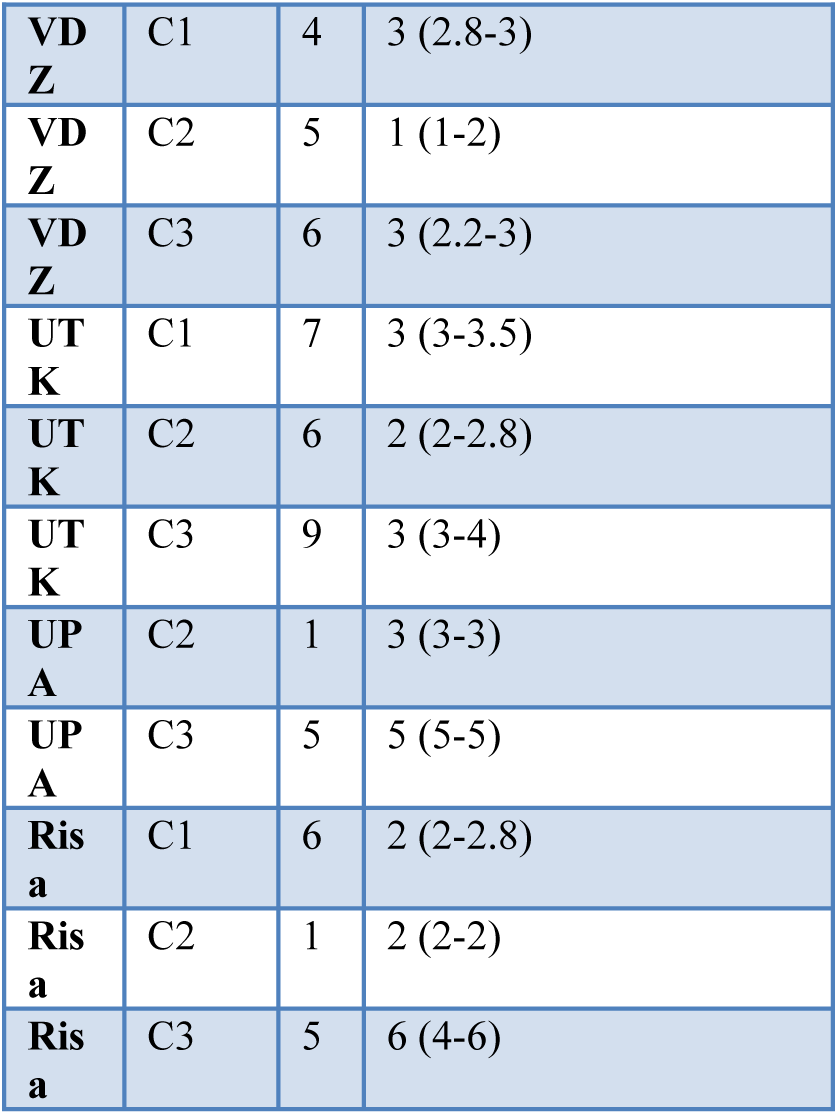
Advanced-therapy line position by drug and cluster.

**Supplementary Table 5.** Advanced-therapy exposure duration by drug and cluster.

| **Dru**<br>**g** | **Clust**<br>**er** | **N** | **Duration, years, median**<br>**(IQR)** |
| --- | --- | --- | --- |
| **IFX** | C1 | 1<br>1 | 1 (0.42-1.29) |
| **IFX** | C2 | 1<br>0 | 1 (1-1.46) |
| **IFX** | C3 | 7 | 0.92 (0.37-2.62) |
| **AD**<br>**A** | C1 | 2<br>2 | 2.75 (0.56-4) |
| **AD**<br>**A** | C2 | 1<br>0 | 2.38 (0.69-3.63) |
| **AD**<br>**A** | C3 | 1<br>6 | 1.13 (0.5-2.37) |
| **VD**<br>**Z** | C1 | 1 | 0.33 (0.33-0.33) |
| **VD**<br>**Z** | C2 | 2 | 0.71 (0.56-0.85) |
| **VD**<br>**Z** | C3 | 4 | 0.54 (0.42-0.69) |
| **UT**<br>**K** | C1 | 2 | 1.29 (1.27-1.31) |
| **UT**<br>**K** | C2 | 2 | 1.29 (0.73-1.85) |
| **UT**<br>**K** | C3 | 3 | 0.5 (0.29-2.46) |
| **UP**<br>**A** | C2 | 1 | 0.92 (0.92-0.92) |
| **UP**<br>**A** | C3 | 3 | 0.67 (0.5-0.83) |
| **Ris**<br>**a** | C3 | 2 | 0.46 (0.44-0.48) |

**Supplementary Table 6.**
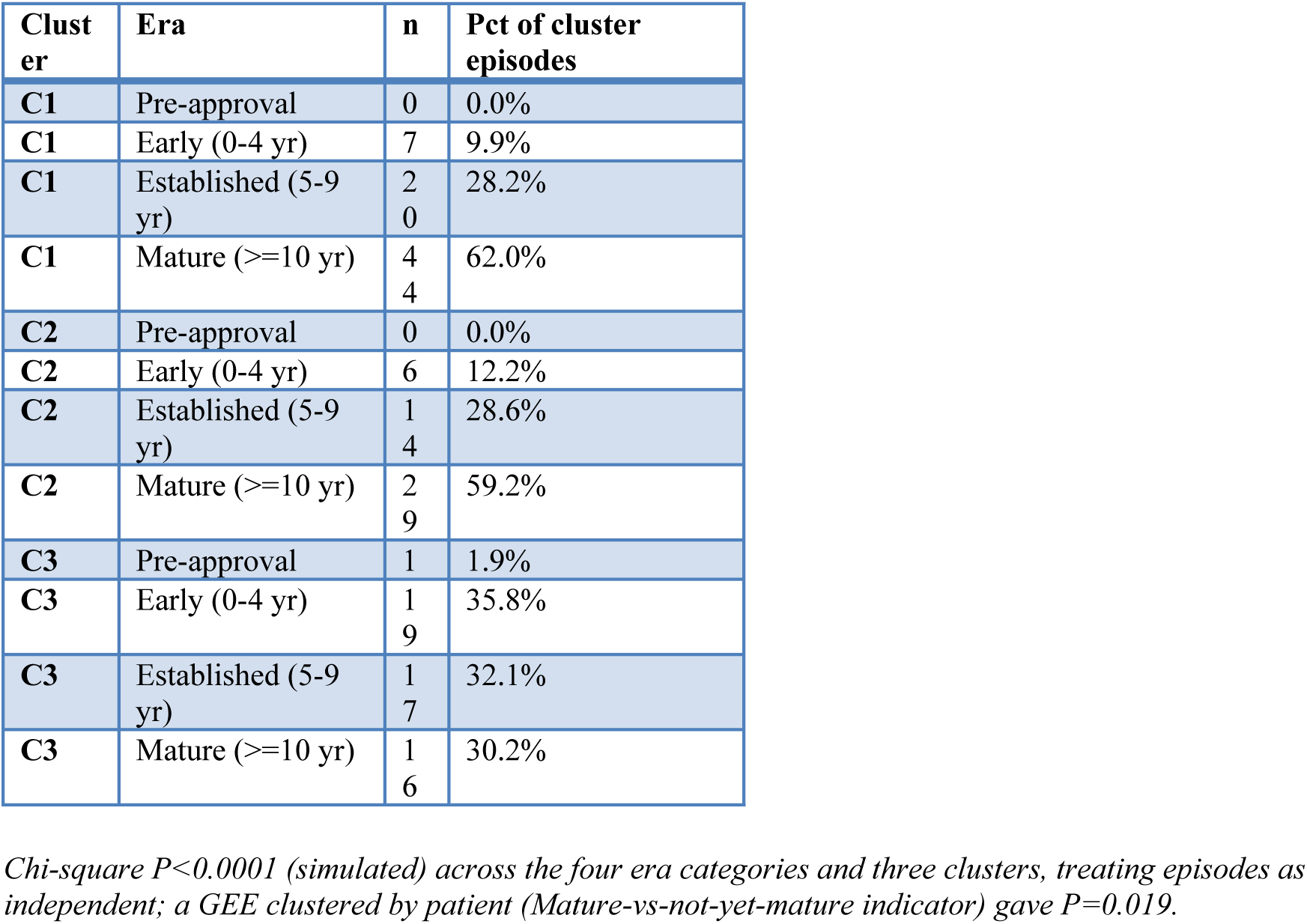
Treatment era distribution by cluster.

**Supplementary Table 7.** Treatment era distribution by cluster and drug.

| Cluster | Drug | Era | n | Pct of cluster-drug episodes |
| --- | --- | --- | --- | --- |
| C1 | Infliximab (IFX) | Pre-approval | 0 | 0.0% |
| C1 | Infliximab (IFX) | Early (0-4 yr) | 1 | 4.5% |
| C1 | Infliximab (IFX) | Established (5-9 yr) | 4 | 18.2% |
| C1 | Infliximab (IFX) | Mature ($\geq 10$ yr) | 17 | 77.3% |
| C1 | Adalimumab (ADA) | Pre-approval | 0 | 0.0% |
| C1 | Adalimumab (ADA) | Early (0-4 yr) | 3 | 7.5% |
| C1 | Adalimumab (ADA) | Established (5-9 yr) | 11 | 27.5% |
| C1 | Adalimumab (ADA) | Mature ($\geq 10$ yr) | 26 | 65.0% |
| C1 | Vedolizumab (VDZ) | Pre-approval | 0 | 0.0% |
| C1 | Vedolizumab (VDZ) | Early (0-4 yr) | 1 | 33.3% |
| C1 | Vedolizumab (VDZ) | Established (5-9 yr) | 1 | 33.3% |
| C1 | Vedolizumab (VDZ) | Mature ($\geq 10$ yr) | 1 | 33.3% |
| C1 | Ustekinumab (UTK) | Pre-approval | 0 | 0.0% |
| C1 | Ustekinumab (UTK) | Early (0-4 yr) | 1 | 20.0% |
| C1 | Ustekinumab (UTK) | Established (5-9 yr) | 4 | 80.0% |
| C1 | Ustekinumab (UTK) | Mature ($\geq 10$ yr) | 0 | 0.0% |
| C1 | Risankizumab (Risa) | Pre-approval | 0 | 0.0% |
| C1 | Risankizumab (Risa) | Early (0-4 yr) | 1 | 100.0% |
| C1 | Risankizumab (Risa) | Established (5-9 yr) | 0 | 0.0% |
| C1 | Risankizumab (Risa) | Mature ($\geq 10$ yr) | 0 | 0.0% |
| C2 | Infliximab (IFX) | Pre-approval | 0 | 0.0% |
| C2 | Infliximab (IFX) | Early (0-4 yr) | 0 | 0.0% |
| C2 | Infliximab (IFX) | Established (5-9 yr) | 0 | 0.0% |
| C2 | Infliximab (IFX) | Mature ($\geq 10$ yr) | 13 | 100.0% |
| C2 | Adalimumab (ADA) | Pre-approval | 0 | 0.0% |
| C2 | Adalimumab (ADA) | Early (0-4 yr) | 2 | 7.7% |
| C2 | Adalimumab (ADA) | Established (5-9 yr) | 8 | 30.8% |
| C2 | Adalimumab (ADA) | Mature ($\geq 10$ yr) | 16 | 61.5% |
| C2 | Vedolizumab (VDZ) | Pre-approval | 0 | 0.0% |
| C2 | Vedolizumab (VDZ) | Early (0-4 yr) | 2 | 66.7% |
| C2 | Vedolizumab (VDZ) | Established (5-9 yr) | 1 | 33.3% |
| C2 | Vedolizumab (VDZ) | Mature ($\geq 10$ yr) | 0 | 0.0% |
| C2 | Ustekinumab (UTK) | Pre-approval | 0 | 0.0% |
| C2 | Ustekinumab (UTK) | Early (0-4 yr) | 0 | 0.0% |
| C2 | Ustekinumab (UTK) | Established (5-9 yr) | 5 | 100.0% |
| C2 | Ustekinumab (UTK) | Mature ($\geq 10$ yr) | 0 | 0.0% |
| C2 | Upadacitinib (UPA) | Pre-approval | 0 | 0.0% |
| C2 | Upadacitinib (UPA) | Early (0-4 yr) | 2 | 100.0% |
| C2 | Upadacitinib (UPA) | Established (5-9 yr) | 0 | 0.0% |
| C2 | Upadacitinib (UPA) | Mature ($\geq 10$ yr) | 0 | 0.0% |
| C3 | Infliximab (IFX) | Pre-approval | 0 | 0.0% |
| C3 | Infliximab (IFX) | Early (0-4 yr) | 1 | 8.3% |
| C3 | Infliximab (IFX) | Established (5-9 yr) | 2 | 16.7% |
| C3 | Infliximab (IFX) | Mature ($\geq 10$ yr) | 9 | 75.0% |
| C3 | Adalimumab (ADA) | Pre-approval | 0 | 0.0% |
| C3 | Adalimumab (ADA) | Early (0-4 yr) | 4 | 21.1% |
| C3 | Adalimumab (ADA) | Established (5-9 yr) | 8 | 42.1% |
| C3 | Adalimumab (ADA) | Mature ($\geq 10$ yr) | 7 | 36.8% |
| C3 | Vedolizumab (VDZ) | Pre-approval | 0 | 0.0% |
| C3 | Vedolizumab (VDZ) | Early (0-4 yr) | 3 | 42.9% |
| C3 | Vedolizumab (VDZ) | Established (5-9 yr) | 4 | 57.1% |
| C3 | Vedolizumab (VDZ) | Mature ($\geq 10$ yr) | 0 | 0.0% |
| C3 | Ustekinumab (UTK) | Pre-approval | 0 | 0.0% |
| C3 | Ustekinumab (UTK) | Early (0-4 yr) | 4 | 57.1% |
| C3 | Ustekinumab (UTK) | Established (5-9 yr) | 3 | 42.9% |
| C3 | Ustekinumab (UTK) | Mature ($\geq 10$ yr) | 0 | 0.0% |
| C3 | Upadacitinib (UPA) | Pre-approval | 0 | 0.0% |
| C3 | Upadacitinib (UPA) | Early (0-4 yr) | 3 | 100.0% |
| C3 | Upadacitinib (UPA) | Established (5-9 yr) | 0 | 0.0% |
| C3 | Upadacitinib (UPA) | Mature ($\geq 10$ yr) | 0 | 0.0% |
| C3 | Risankizumab (Risa) | Pre-approval | 1 | 20.0% |
| C3 | Risankizumab (Risa) | Early (0-4 yr) | 4 | 80.0% |
| C3 | Risankizumab (Risa) | Established (5-9 yr) | 0 | 0.0% |
| C3 | Risankizumab (Risa) | Mature ($\geq 10$ yr) | 0 | 0.0% |

*N and percentage are calculated within each cluster-drug pair (percentages sum to 100% across the four era categories for a given cluster and drug, not across drugs). Drug-cluster combinations with zero recorded episodes are omitted.*

**Supplementary Table 8.**
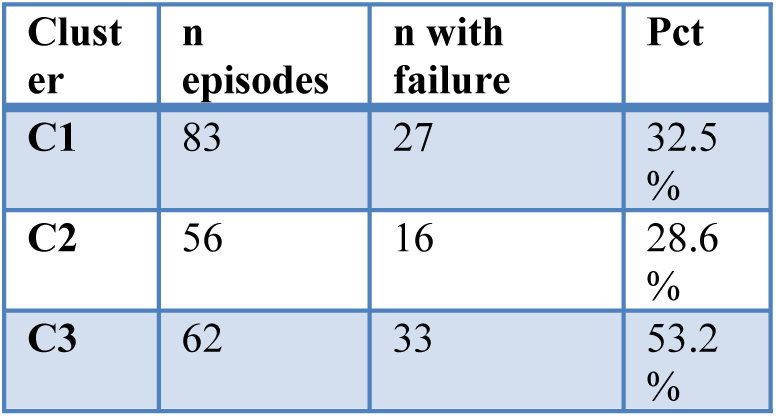
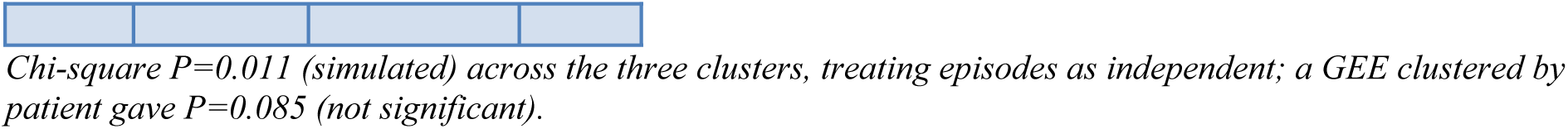
Episodes with documented failure by cluster.

| Cluster | n episodes | n with failure | Pct |
| --- | --- | --- | --- |
| C1 | 83 | 27 | 32.5 % |
| C2 | 56 | 16 | 28.6 % |
| C3 | 62 | 33 | 53.2 % |

**Supplementary Table 9.** Episodes with documented failure by drug, pooled across clusters.

| Drug | n episodes | n with failure | Pct |
| --- | --- | --- | --- |
| IFX | 48 | 24 | 50.0 % |
| ADA | 89 | 33 | 37.1 % |
| VDZ | 18 | 9 | 50.0 % |
| UTK | 26 | 8 | 30.8 % |
| UPA | 7 | 2 | 28.6 % |
| Risa | 13 | 0 | 0.0% |

Chi-square $P=0.022$ (simulated) across the six agents.

**Supplementary Table 10.**
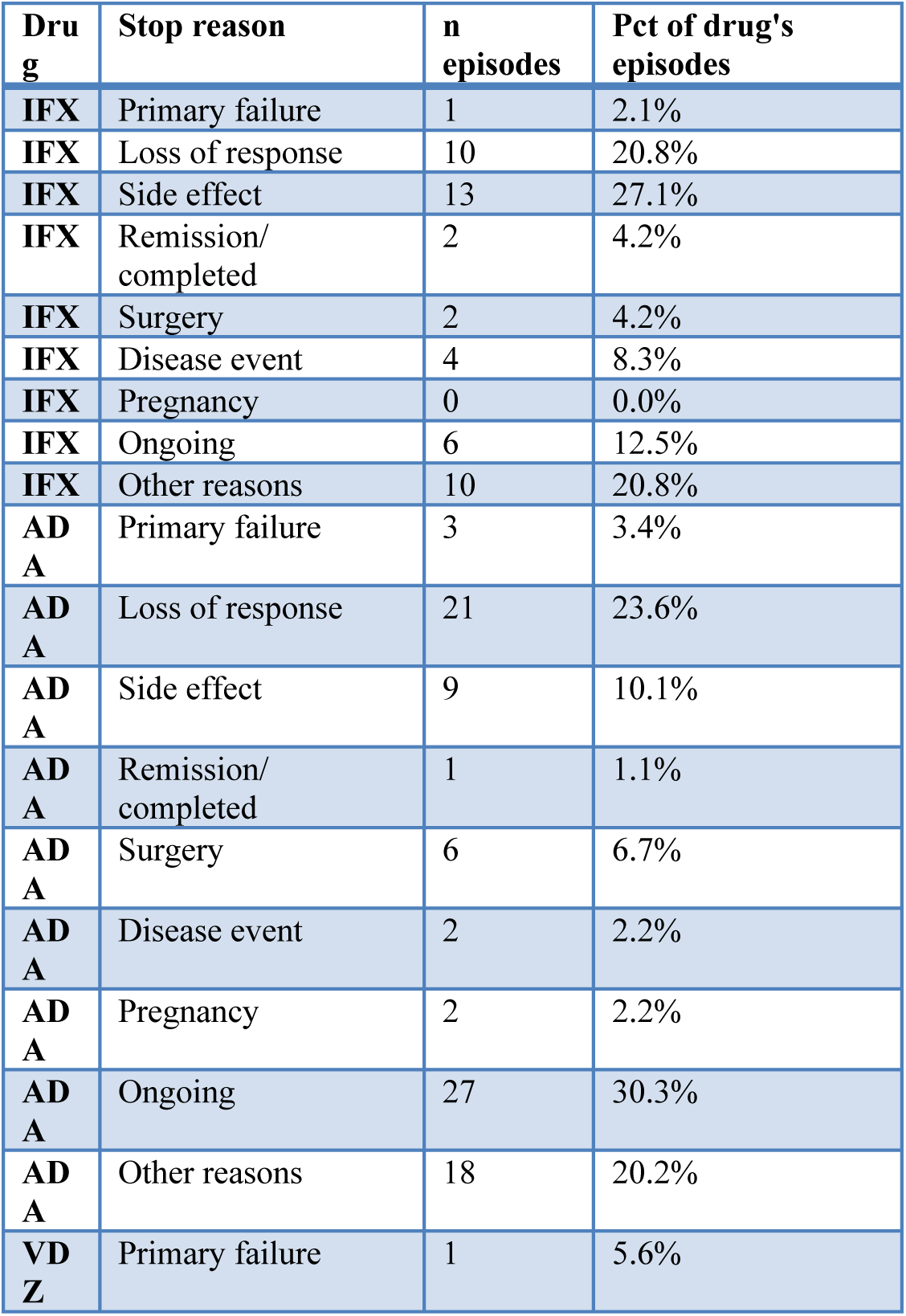
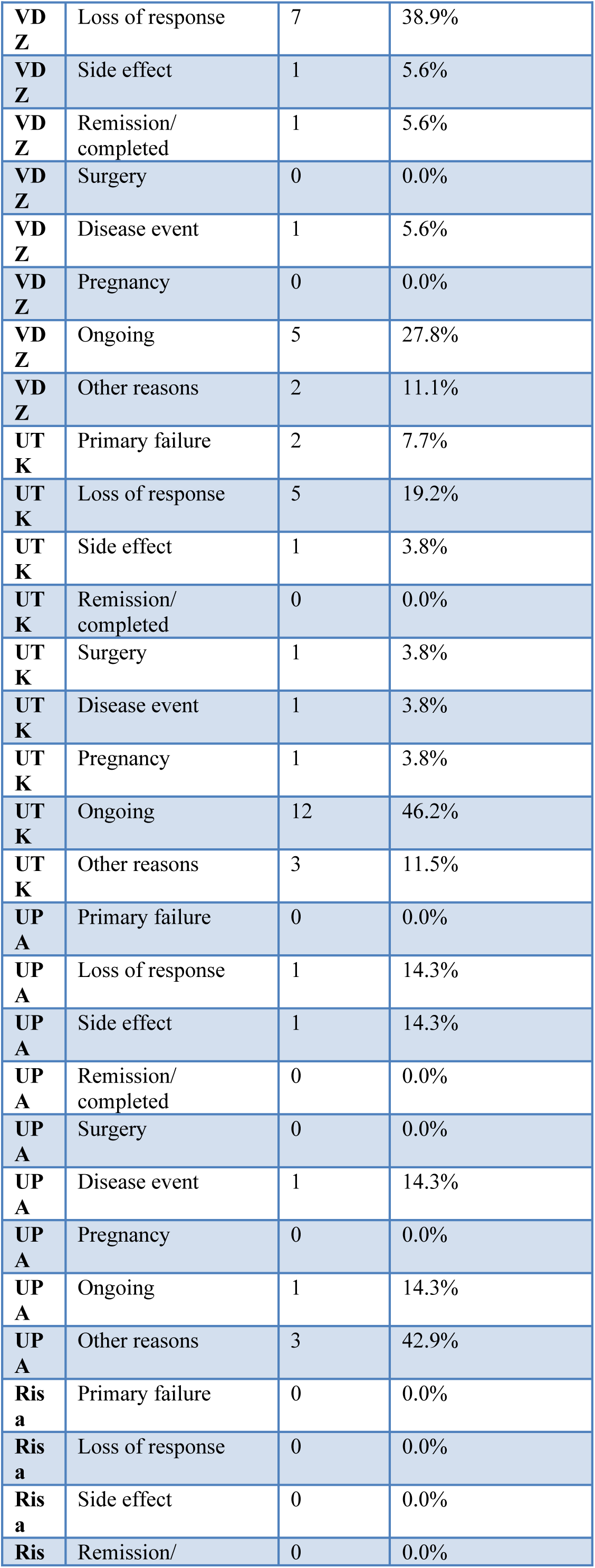
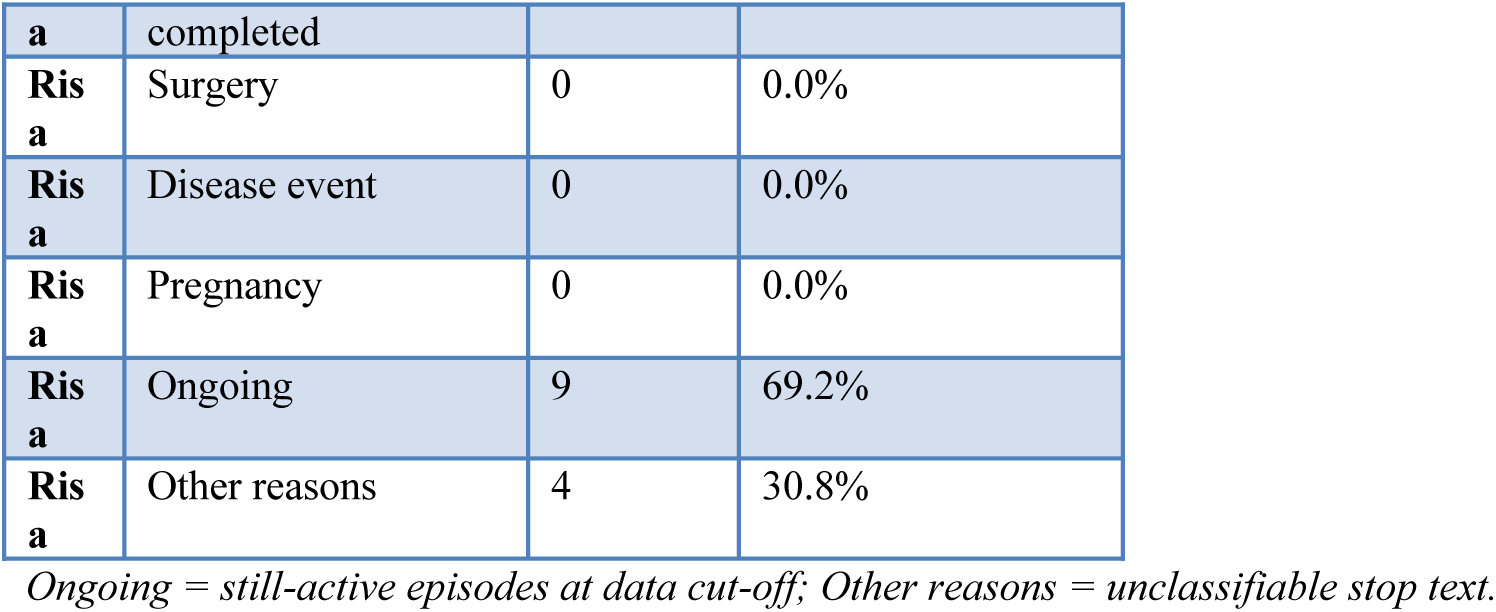
Reason for discontinuation by advanced-therapy agent.

**Supplementary Table 11.**
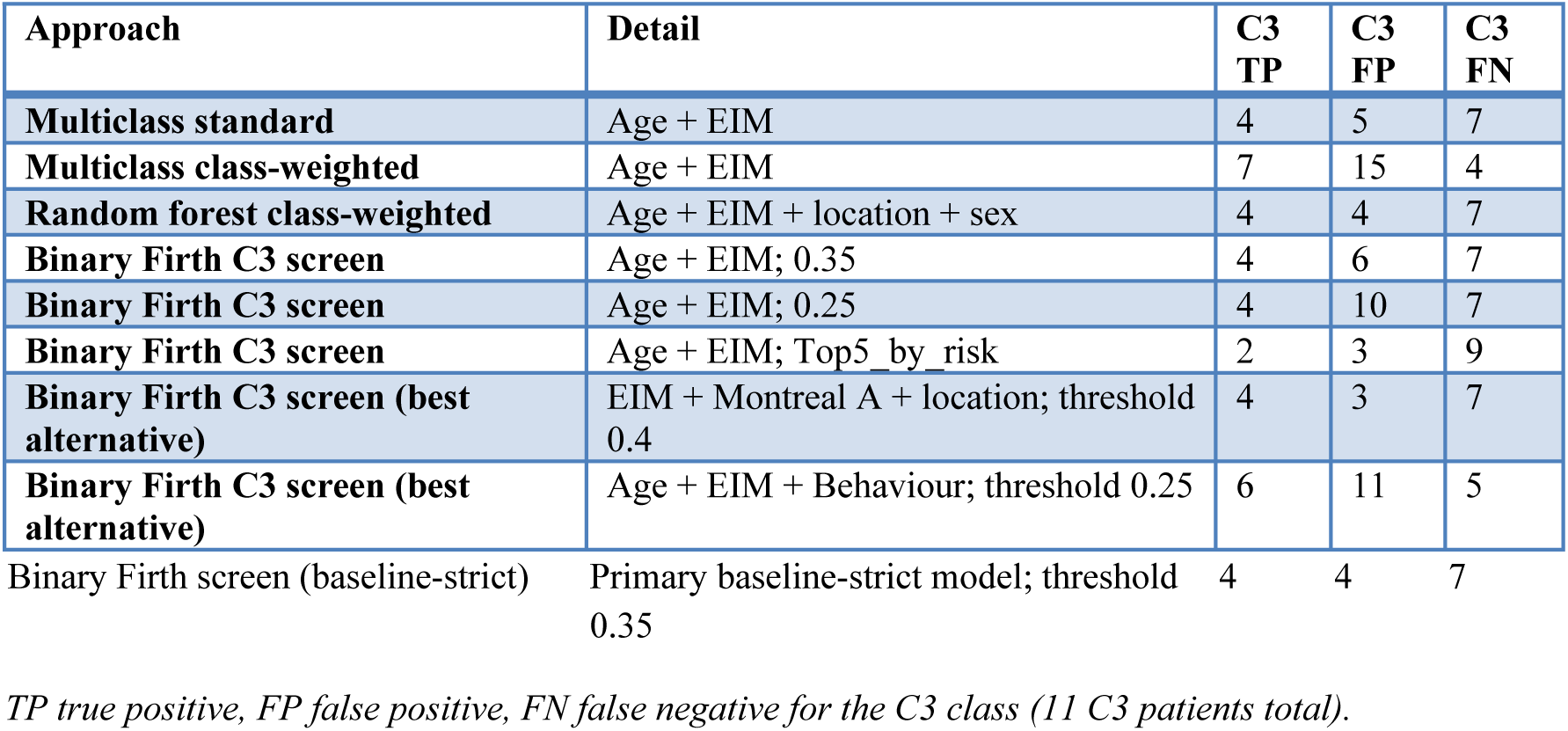
Internal C3-detection trade-off across candidate screening approaches.

**Supplementary Table 12.** Primary baseline model: predicted C3 probability by observed cluster.

| Cluster | N | Mean predicted C3 probability | Median | Min | Max |
| --- | --- | --- | --- | --- | --- |
| **C1** | 56 | 0.097 | 0.067 | 0.013 | 0.676 |
| **C2** | 31 | 0.069 | 0.059 | 0.009 | 0.265 |
| **C3** | 11 | 0.255 | 0.173 | 0.051 | 0.907 |

